# Cytokine Gene Variants Are Associated with the Inflammatory and Metabolic Profile of Human Exercise Performance

**DOI:** 10.1101/2025.11.05.25339580

**Authors:** Kinga Humińska-Lisowska, Barkın Bıçakçı, Monika Michałowska-Sawczyn, Piotr Aschenbrenner, Alison V September, Patrizia Proia, Agata Leońska-Duniec

## Abstract

**Objectives:** Genetic variation in cytokine genes may influence inflammation, metabolism, and exercise-related adaptations, yet large-scale population data remain scarce. This study examined whether five polymorphisms in interleukin-6 (*IL6*; rs1800795, rs1800796, rs1800797), interleukin-15 (*IL15*; rs1589241), and tumour necrosis factor-alpha (*TNF-α*; rs1800629) are associated with physiological, biochemical, and performance-related traits in healthy adults.

**Methods:** A total of 1,000 healthy adults of European ancestry underwent comprehensive physiological, biochemical, and genomic assessments. Participants were classified according to objectively measured maximal oxygen uptake (VO2max), providing a quantitative marker of aerobic capacity rather than athlete-control contrasts.

**Results:** Carriers of the *IL6* rs1800795 CC genotype showed higher serum iron (p = 0.004) and hematocrit (p = 0.033). The *IL6* C-G-G haplotype (rs1800795-rs1800796-rs1800797) was also associated with higher iron levels (p = 0.0012). For *TNF-α* rs1800629, AG and AA genotypes were less likely to belong to the higher VO2max group (p = 0.006). AG carriers also had nominally higher LDL cholesterol (p = 0.037), while AA carriers showed a trend toward a longer time to peak power in the Wingate test (p = 0.052). No significant single-locus effects were detected for *IL15*, but the *IL15* CC/CT × *TNF-α* GG combination was associated with greater odds of higher aerobic capacity (p = 0.004).

**Conclusion:** These findings reveal modest but consistent genetic influences on iron metabolism, lipid profile, and aerobic capacity. Variants in *IL6* and *TNF-α* were linked to performance-relevant traits, while a multi-locus *IL15* × *TNF-α* interaction further supports the role of cytokine-related gene networks in individual differences in aerobic fitness and recovery potential.

**WHAT IS ALREADY KNOWN ON THIS TOPIC:**

- Genes that regulate inflammation, such as interleukin-6 (*IL6*), interleukin-15 (*IL15*) and tumour necrosis factor-alpha (*TNF-α*), may influence how people respond to exercise and recover after training.
- Prior studies were often small and compared athletes with non-athletes, rather than using an objective physiological classification like VO2max-leaving population-level links uncertain.

**WHAT THIS STUDY ADDS:**

- In a study of 1,000 healthy European adults, *IL6* variants were linked with higher iron and hematocrit - factors that can support oxygen transport and recovery.
- People carrying the *TNF-α* A variant tended to have lower aerobic capacity, while a specific combination of *IL15* and *TNF-α* variants was linked to better aerobic fitness (higher VO2max).

**HOW THIS STUDY MIGHT AFFECT RESEARCH, PRACTICE OR POLICY:**

- These results suggest that differences in immune-related genes partly explain why people respond differently to training.
- VO2max-based genetic profiling could support more tailored training, recovery, and prevention strategies in sport and clinical practice.

## 1. INTRODUCTION

The positive impact of physical activity on reduced mortality, decreased risk of developing non-communicable diseases, and improved mental health has been well recognized for decades [1,2]. Despite this, the mechanisms that translate repeated bouts of exercise into durable adaptations are not fully understood. A central working model posits an inflammatory- regenerative axis in which acute, contraction-induced signals trigger systemic communication that drives remodelling and performance improvements. However, searching for a muscle contraction-induced humoral factor, an “exercise factor” that could cause some of the exercise-induced changes in major metabolic organs represents a novel challenge. Recent research has revealed that skeletal muscle in response to external stimuli can secrete a group of biochemical molecules usually known as myokines [3,4]. In general, myokines are cytokines and other peptides that are produced, expressed, and released by muscle fibres and regulate whole-body metabolism in an autocrine, paracrine, or endocrine manner [4,5]. The conceptual basis of considering these products as key mediators within the complex organ communication network is essential for developing a novel understanding of the molecular basis of physical activity [6]. This framework is in line with emerging evidence that biomarkers of tissue stress and inflammation can be used to monitor training load and recovery.

Although it had long been suggested, it was ∼25 years ago that the first proposed muscle contraction-induced factor mediating exercise effects in other organs, interleukin-6 (IL6), was described by Pedersen and coworkers [7]. Ongoing research demonstrates that skeletal muscle tissue may produce inflammatory biomarkers belonging to different families such as interleukin-15 (IL15) or tumor necrosis factor-alpha (TNF-α) [4,8,9]. These interleukins potentially have anabolic effects in human skeletal muscle, can be associated with hypertrophy muscle growth, play important role in regulating energy metabolism, and consequently training adaptation [4,10]. Convincing evidence exists that the expression of IL6 and IL15 is regulated by a bout of exercise [8,11]. In addition, IL6 expression during muscle contraction may inhibit the production of the proinflammatory cytokine TNF-α, which is significantly involved, among others, in glucose metabolism [9]. Our group has also demonstrated that systemic biomarkers (cfDNA, adipokines) respond to exercise intensity and body composition, supporting the relevance of inflammation-related signals in training [12,13]. Taken together, IL6, IL15, and TNF-α form a biologically plausible and tractable triad through which the inflammatory- regenerative axis influences phenotype and performance. However, interindividual variability in myokine production and release has been observed due to functional single nucleotide polymorphisms (SNPs) in the genes encoding these molecules. These genetic variants may affect gene transcription and cytokine synthesis, altering the intensity of the inflammatory response, individual susceptibility to certain diseases, as well as effectiveness of exercise training programs [14]. Therefore, genetic modulation of myokine signaling is a plausible cause of the differences between people in how they adapt to training stimuli.

Although almost 50 SNPs have been described for *IL6* gene (7p15.3) so far [15], the SNPs located in the promoter region seem to be particularly important because they may quantitatively change the gene’s expression [16]. Thus, three promoter *IL6* polymorphisms, namely, rs1800795 (-174 G > C), rs1800796 (-572 G > C), and rs1800797 (-597 A > G), associated with *IL6* transcription activity, were selected for this study. In addition, to get a better picture of the complex interactions among various genetic variants, SNPs rs1589241 (T>A) in the regulatory element (first intron, 3’UTR) of *IL15* (4q31.21) and rs1800629 (-308 G>A) in the promoter region of *TNF-α* (6p21.33) were added to the analysis. It is noteworthy that only simultaneous analysis of numerous polymorphic sites can provide additional unique information about the associations between genetic background and phenotypic traits, as well as insight into the dependency among genetic markers [17]. Therefore, we prioritized a parsimonious and biologically grounded candidate panel spanning myokine production (IL6 and IL15) and proinflammatory tone (TNF-α).

Given that genetic variations may affect cytokine production, we hypothesized that polymorphic sites in the *IL6*, *IL15*, and *TNF-α* genes could interact with inflammatory markers, body composition, basic biochemical parameters, and variables of aerobic and anaerobic performance. The present study therefore aimed to investigate both the individual and combined roles of five SNPs - three in the *IL6* promoter region (rs1800795, rs1800796, and rs1800797), one in *IL15* (rs1589241) and one in *TNF*-α (rs1800629) - in physically active Polish individuals aged 20-57 with varying performance levels. Specifically, we examined whether these SNPs are associated with differences in body composition, selected blood parameters, and both aerobic and anaerobic exercise capacity. Because these proinflammatory and myokine-related genes may underlie individual variability in training response and exercise adaptation, we further explored potential interactions among these variants (e.g., *IL6* haplotypes combined with *IL15* and *TNF-α* genotypes) to determine whether specific genetic profiles coincide with the observed variation in muscle-related traits and metabolic indices. This cross-sectional study is part of a larger research program that examines the inflammatory-regenerative axis of exercise adaptation using circulating biomarkers, microbiome features, and genetics. This study focuses specifically on the genetic component of this framework.

## 2. MATERIALS AND METHODS

### 2.1 Study design and participants

This cross-sectional study was part of the “1000 Genomes – genetic basis of physical activity, sport level and well-being” program at the University of Physical Education and Sport in Gdańsk. Of the 1,000 adults of self-reported Polish ancestry who were recruited, 964 were enrolled. The analyses included only individuals with complete maximal oxygen uptake (VO2max), cardiovascular/haematological, and genotyping data.

The inclusion criteria were as follows: age between 20 and 57 years, good general health, absence of a diagnosed cardiovascular, metabolic, autoimmune, or neurological disease, no current use of anti-inflammatory, hormonal, or performance-enhancing medications, written informed consent, completion of standardized physiological testing including VO2max, and provision of venous blood, buccal swab, and stool samples. The exclusion criteria were missing VO2max, use of drugs that affect inflammatory, endocrine, or cardiometabolic function, acute injury or infection within the previous three months, and genetic data that failed quality control. Participants attended a cooperating hospital in a fasted state for screening (fasting blood and urine tests, chest X-ray). Eligible individuals received home sampling kits for saliva, stool, oral swabs, and urine, and were scheduled for laboratory assessments. Each participant was assigned an anonymized study code. The study complied with the Declaration of Helsinki and was approved by the Bioethics Committee at the District Medical Chamber in Gdańsk (KB-16/20). Written informed consent was obtained for biospecimen collection, genetic analyses, and use of anonymized data.

### 2.2 Anthropometric, body composition, and performance testing

Anthropometry and body composition were assessed under standardized conditions. Stature was measured to 0.1 cm with a wall stadiometer and body mass was measured to 0.1 kg with a calibrated digital scale. Body composition, including fat mass, fat-free mass, and total body water, was obtained by multi-frequency bioelectrical impedance analysis using an InBody 720 (Biospace). Waist and hip circumferences were measured with a non-elastic tape according to the World Health Organization protocol. Body mass index (BMI) was calculated as weight divided by height squared (kg/m²). Testing occurred in a fasted state or at least three hours after a light meal, consistent with bioimpedance guidelines.

Aerobic performance was evaluated using an incremental cycling test to volitional exhaustion with breath-by-breath gas analysis (Jaeger Oxycon Pro, CareFusion) on an electronically braked ergometer (Ergoline Ergoselect viasprint 150p). After a five-minute warm-up at 60 revolutions per minute with a load of 1.0 W·kg^-1^, the protocol consisted of two-minute stages, beginning at 1.5 W·kg^-1^ with 0.5 W·kg^-1^ increments per stage, until exhaustion. Oxygen uptake (VO2), carbon dioxide output (VCO2), respiratory exchange ratio (RER), and minute ventilation (VE) were recorded continuously. Heart rate (HR) was monitored using a Polar H9 chest strap.

Anaerobic performance was evaluated using a 30-second Wingate test on a friction-loaded cycle ergometer (Monark 894E). The braking force was set at 0.075 kilograms per kilogram of body mass. Peak and mean power were expressed relative to body mass (W·kg^-1^).

### 2.3 Biochemical and haematological analyses

Trained personnel at the cooperating hospital (7th Navy Hospital in Gdańsk) drew fasting venous blood into EDTA and serum tubes and processed it according to standard operating procedures. Analyses were performed at a certified clinical laboratory (Diagnostyka Sp. z o.o.) using automated analysers.

A complete blood count includes the following: white blood cell count (WBC), red blood cell count (RBC), haemoglobin (HGB), haematocrit (HCT), mean corpuscular volume (MCV), mean corpuscular haemoglobin (MCH), mean corpuscular haemoglobin concentration (MCHC), and platelets (PLT).

Serum biochemistry: glucose, creatinine, iron, cortisol, total cholesterol (TC), high-density lipoprotein (HDL), low-density lipoprotein (LDL), and triglycerides (TG).

### 2.4 DNA isolation, genotyping, and statistical analysis

Genomic deoxyribonucleic acid (DNA) was extracted from buccal epithelial cells collected with sterile FLOQSwabs (Copan) using the High Pure PCR Template Preparation Kit (Roche), following the manufacturer’s instructions. The genotyping targeted the following variants: interleukin-6 (IL6) promoter variants rs1800795 (−174G>C, assay ID C 1839697_20), rs1800796 (−572G>C, C 11326893_10), and rs1800797 (−597A>G, C 1839695_20); interleukin-15 (IL15) variant rs1589241 (C 8865652_10); and tumour necrosis factor-alpha (TNF-α) variant rs1800629 (−308G>A, C 7514879_10). Allelic discrimination was performed using TaqMan® SNP Genotyping Assays (Applied Biosystems) on a real-time polymerase chain reaction (PCR) platform (Bio-Rad CFX96).

Each 5-μL reaction contained the following: 2.5 μL of TaqPath™ ProAmp™ Master Mix; 0.25 μL of a 10× SNP assay mix containing VIC/FAM-labelled probes; 1.0 μL of nuclease-free water; and 1.0 μL of genomic DNA. The thermocycling protocol included a pre-read at 60 °C for 30 seconds, an initial denaturation at 95 °C for five minutes, and 40 cycles of denaturation at 95 °C for five seconds and annealing/extension at 60 °C for 30 seconds. A post-read at 60 °C for 30 seconds was also included. Fluorescence clusters were visualized and genotype calls were reviewed in CFX Maestro software (Bio-Rad). Quality control included sample and marker call rate thresholds, duplicate checks, and Hardy-Weinberg equilibrium testing by an exact test in the pooled sample and within fitness categories.

### 2.5 Statistical analysis

Group differences by genotype were examined using Kruskal-Wallis rank sum test. Linear models and analysis of covariance (ANCOVA) were adjusted for age and body mass. In single-locus analysis predefined inheritance models (codominant, dominant, recessive, overdominant, and log-additive). Hardy-Weinberg equilibrium and association analyses were performed in R [18] using the SNPassoc package (version 2.1-2). Results are presented with effect estimates and 95% CIs where applicable, and two-sided P values. Statistical reporting adhered to the CHAMP checklist for transparency and rigor (eg, model specification, variable coding, assumption checks, handling of small cell counts), with the CHAMP statement [19].

#### 2.5.1 IL-6 haplotype-based analysis

Haplotype-based *IL6* association analyses were conducted in two steps. A prespecified subset of phenotypes that showed association at the single-locus levels were tested using a hierarchical approach: an omnibus *IL6* haplotype test for each phenotype (using haplo.score), and for phenotypes with significant global evidence, haplotype-specific regression models (using haplo.glm function) to obtain effect estimates (adjusted for age, sex). Multiple testing across primary phenotypes was controlled using the Benjamini–Hochberg false discovery rate (FDR) applied to omnibus P values, within any significant phenotype, haplotype-specific p-values were FDR-adjusted. To screen remaining phenotypes, we repeated the omnibus *IL6* haplotype test. These results were error controlled using BH-FDR across all omnibus tests in this step. Follow-up haplotype-specific effects were only summarized for exploratory phenotypes passing the FDR threshold. Haplo.score and haplo.glm functions are available in the haplo.stats R package (version 1.9.7)

#### 2.5.2 Targeted gene-gene interaction

Targeted gene-gene interaction analyses were pre-specified for the union of phenotypes showing single-locus signals with *IL6*, *IL15* and *TNF-a*. For each selected phenotype we added a block of *IL6*×*IL15* (and separately *IL6*×*TNF-α*) interaction terms for the most frequent (>1% for continuous, and >10% for VO2max) non-reference *IL6* haplotypes (haplo.glm) and compared models with and without these terms using block likelihood-ratio tests. Multiple testing was controlled using the Benjamini–Hochberg FDR (Q value threshold 0.100), while the nominal significance was set at P < 0.05.

#### 2.5.3 Data-driven model free approach

To analyse higher-order and non-additive interactions among gene variants, we applied a logic regression framework integrating SNPs from the *IL6*, *IL15*, and *TNF* loci. Logic regression is a model-free, data-driven approach that identifies combinations of binary predictors that best explain categorical outcome. The analysis was conducted using the LogicFS R package (version 2.22.0). Each SNP was represented by two binary dummy variables to capture all three genotypic states: SNP_1 = 0, SNP_2 = 0 for homozygous reference, SNP_1 = 1, SNP_2 = 0 for heterozygous; and SNP_1 = 1, SNP_2 = 1 for homozygous variant. To prevent overfitting and for evaluation purposes, we performed 100 bootstrap runs on real data and 100 runs on phenotype-resampled (VO2max status shuffled) data to obtain null distributions. For each run, importance scores were computed to quantify the contribution of individual genotype combinations to model performance. Combinations that occurred in more than 10% of real-data iterations were retained for further evaluation. Bootstrap-based 95% confidence intervals for importance scores were estimated to assess stability and significance relative to the resampled data.

### 2.6 Equity, diversity, and inclusion statement

This study was designed and conducted in alignment with the principles of equity, diversity, and inclusion (EDI). We recruited adults from across Poland (multiple regions and urban/rural settings) and aimed for balance by sex and age within the eligibility constraints of a healthy, physically active cohort. Study visits were scheduled to accommodate work/study hours, with clear plain-language materials and standardized procedures to support inclusive participation. Analyses included sex- and age-adjusted models and we report sex distribution and key demographics; we do not generalize beyond community-dwelling European adults and acknowledge this as a limitation. The multidisciplinary author team includes investigators at various academic levels (junior and senior researchers) and reflects collaboration across physiology, sports medicine, genetics, and biostatistics.

## 3. RESULTS

Participants were assigned to seven predefined VO2max categories by sex and age (adapted from Shvartz and Reibold [20], Table 1_SupplInfo). For the analyses, categories were dichotomized into High Fitness (HF, very good, outstanding, N = 311) and Low Fitness (LF, very poor, poor, satisfactory, N = 190). Those classified as medium or good (N = 359) were excluded. All traits stratified by the fitness are presented in Table 1.

**Table 1.**
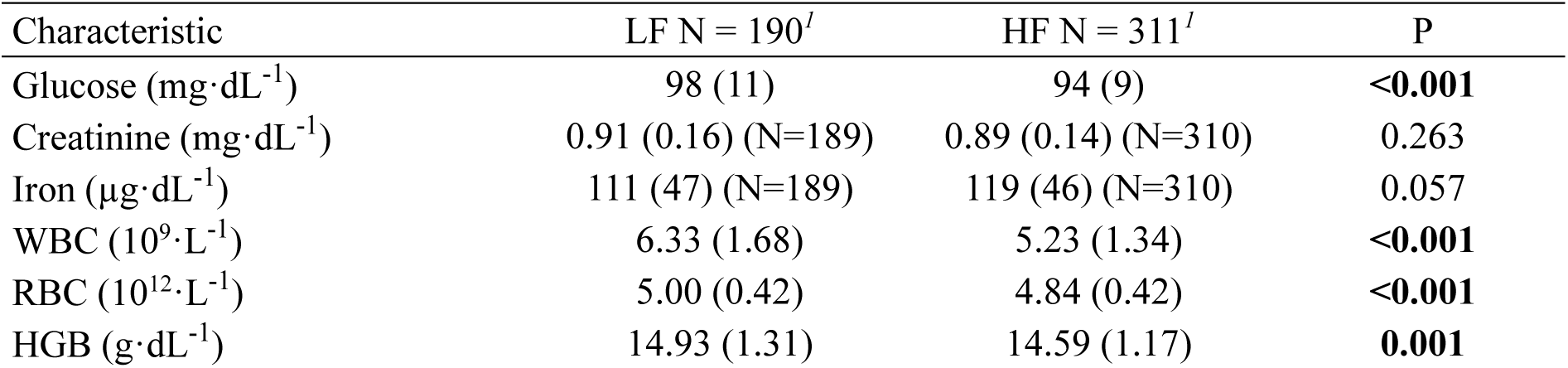
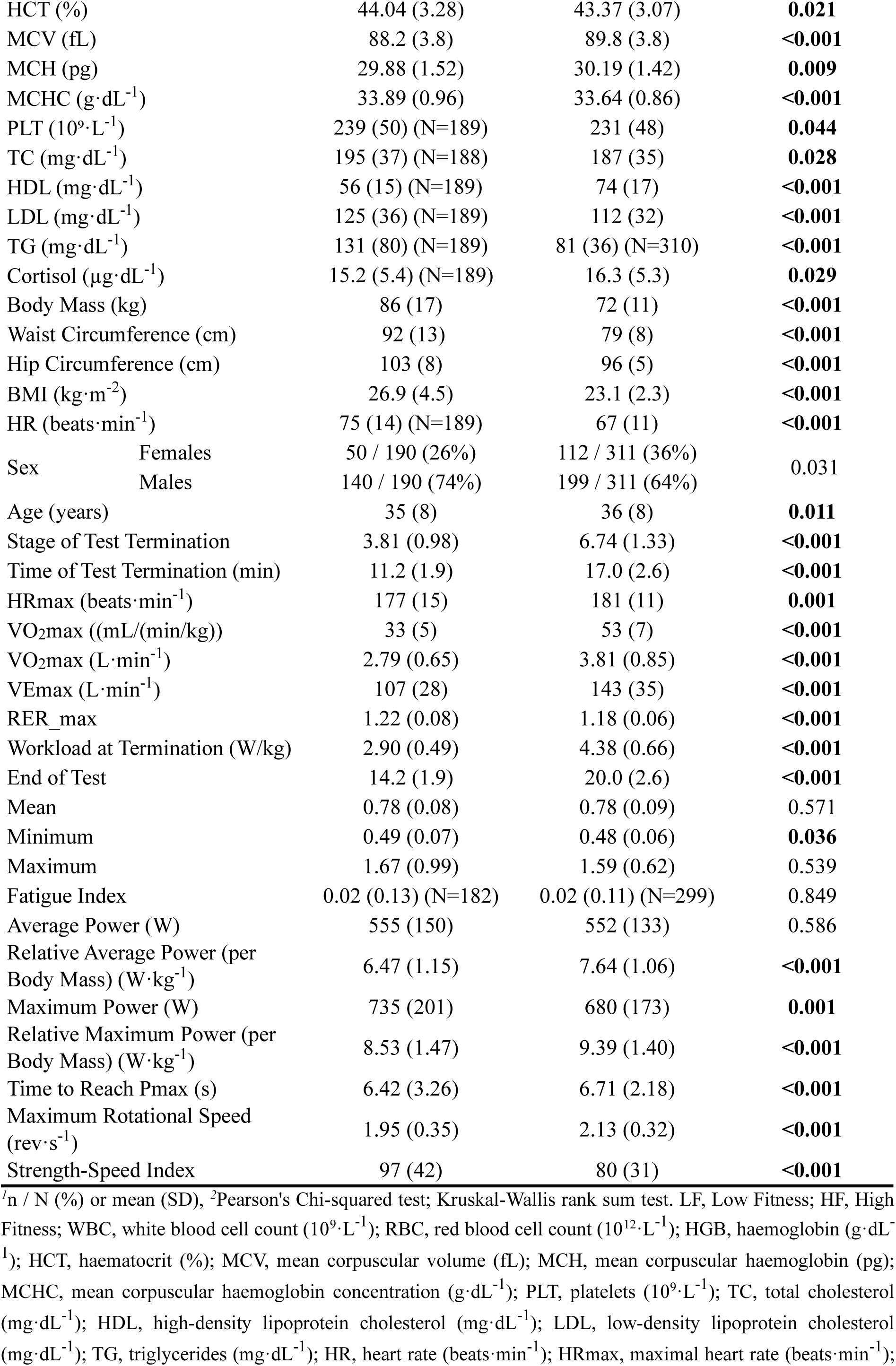
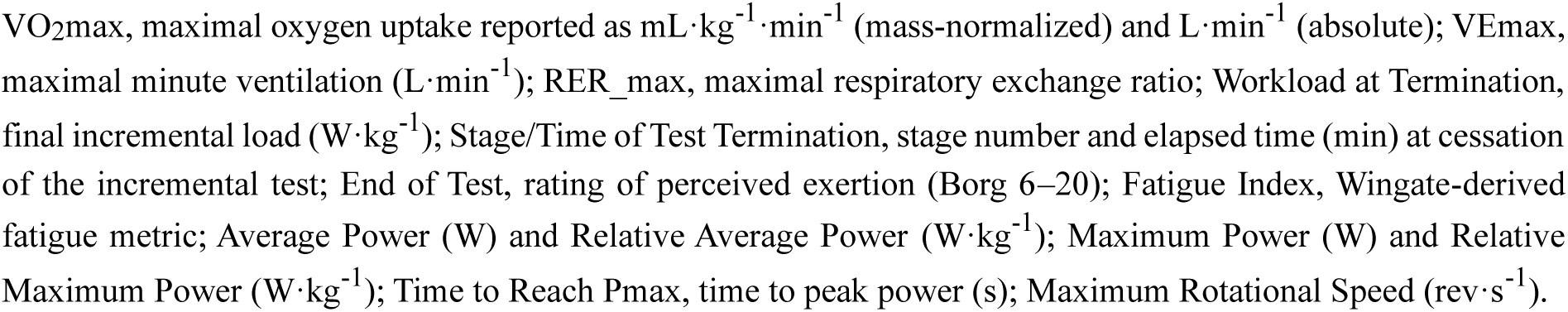
Baseline biochemical, hematological, anthropometric, and performance characteristics by aerobic fitness group (Low Fitness, LF vs High Fitness, HF).

All SNPs conformed to HWE within LF and HF groups except *TNFα* rs1800629 in HF (P=0.034). In the pooled sample, *IL15* rs1589241 (P=0.022) and *IL6* rs1800797 (P=0.00018) deviated from HWE (Table 2_SupplInfo).

### 3.1 Single locus-analysis

Across the assessed domains - body composition (bioimpedance: skeletal muscle mass, fat mass, body-fat percentage), blood parameters (CBC, lipid profile, glucose, creatinine, iron, cortisol), and performance tests (VO2max on cycle ergometer; Wingate peak and mean power) - we identified several unadjusted (nominal) genotype - phenotype associations. Specifically, *IL6* rs1800795 was associated with serum iron (P = 0.004), and haematocrit (P = 0.033); and *TNFα* rs1800629 with fitness group (LF vs. HF; P = 0.006), and LDL cholesterol (P = 0.037) (Supplementary Tables 3-7). *IL15* rs1589241 did not show any nominal associations within the final set of outcomes. These signals were subsequently examined under predefined genetic inheritance models (Tables 3-7). For serum iron (*IL6* rs1800795), associations remained significant under all models except recessive (codominant: P=0.006, FDR-adjusted [Q]=0.015, dominant: P=0.002, Q=0.010, overdominant: P=0.009, Q=0.015, log-additive: P=0.029, Q=0.036, recessive: P=0.783, Q=0.783, Table 3).

**Table 3.**
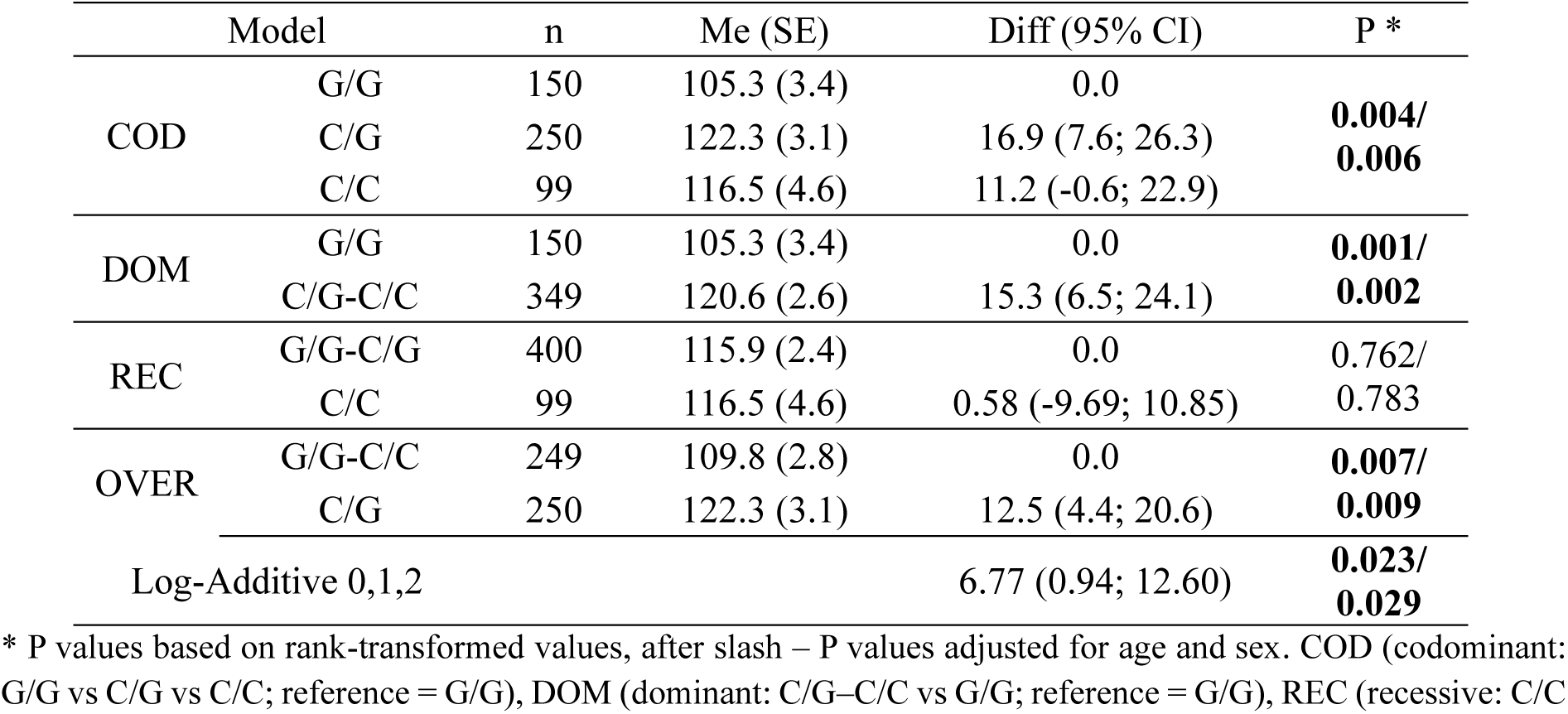
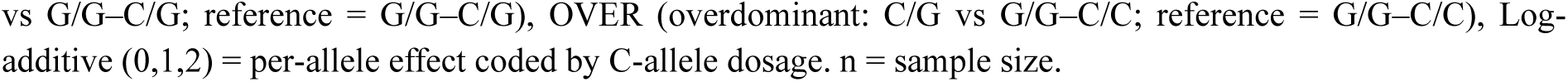
Association of *IL6* rs1800795 with serum iron (µg·dL^-1^)

|  | Model | n | Me (SE) | Diff (95% CI) | P * |
| --- | --- | --- | --- | --- | --- |
| COD | G/G | 150 | 105.3 (3.4) | 0.0 | **0.004/**<br>**0.006** |
|  | C/G | 250 | 122.3 (3.1) | 16.9 (7.6; 26.3) |  |
|  | C/C | 99 | 116.5 (4.6) | 11.2 (-0.6; 22.9) |  |
| DOM | G/G | 150 | 105.3 (3.4) | 0.0 | **0.001/**<br>**0.002** |
|  | C/G-C/C | 349 | 120.6 (2.6) | 15.3 (6.5; 24.1) |  |
| REC | G/G-C/G | 400 | 115.9 (2.4) | 0.0 | 0.762/<br>0.783 |
|  | C/C | 99 | 116.5 (4.6) | 0.58 (-9.69; 10.85) |  |
| OVER | G/G-C/C | 249 | 109.8 (2.8) | 0.0 | **0.007/**<br>**0.009** |
|  | C/G | 250 | 122.3 (3.1) | 12.5 (4.4; 20.6) |  |
| Log-Additive 0,1,2 |  |  |  | 6.77 (0.94; 12.60) | **0.023/**<br>**0.029** |

\* P values based on rank-transformed values, after slash – P values adjusted for age and sex. COD (codominant: G/G vs C/G vs C/C; reference = G/G), DOM (dominant: C/G–C/C vs G/G; reference = G/G), REC (recessive: C/C
vs G/G–C/G; reference = G/G–C/G), OVER (overdominant: C/G vs G/G–C/C; reference = G/G–C/C), Log-additive (0,1,2) = per-allele effect coded by C-allele dosage. n = sample size.

After multiple-testing correction, only LDL cholesterol remained associated with TNFα rs1800629 showed significant associations (codominant model: P=0.029, Q=0.065, overdominant model: P=0.011, Q=0.055, Table 5), while the fitness level was not significant in any models once FDR adjustment was applied (Table 6).

**Table 4.** Association of *IL6* rs1800795 with haematocrit (HCT, %)

|  | Model | n | Me (SE) | Dif (95% CI) | P* |
| --- | --- | --- | --- | --- | --- |
| COD | G/G | 151 | 43.04 (0.26) | 0.0 | **0.033/**<br>0.063 |
|  | C/G | 250 | 43.96 (0.19) | 0.92 (0.29; 1.56) |  |
|  | C/C | 100 | 43.69 (0.35) | 0.65 (-0.14; 1.44) |  |
| DOM | G/G | 151 | 43.04 (0.26) | 0.0 | **0.0096/**<br>**0.020** |
|  | C/G–C/C | 350 | 43.88 (0.17) | 0.84 (0.24; 1.44) |  |
| REC | G/G–C/G | 401 | 43.61 (0.15) | 0.0 | 0.604/<br>0.274 |
|  | C/C | 100 | 43.69 (0.35) | 0.08 (-0.62; 0.77) |  |
| OVER | G/G–C/C | 251 | 43.29 (0.21) | 0.0 | 0.050/<br>0.211 |
|  | C/G | 250 | 43.96 (0.19) | 0.66 (0.11; 1.21) |  |
| log-Additive 0,1,2 |  |  |  | 0.39 (-0.01; 0.78) | **0.046/**<br>**0.032** |

\* P values based on rank-transformed values. COD (codominant: G/G vs C/G vs C/C; reference = G/G), DOM (dominant: C/G–C/C vs G/G; reference = G/G), REC (recessive: C/C vs G/G–C/G; reference = G/G–C/G), OVER (overdominant: C/G vs G/G–C/C; reference = G/G–C/C), log-additive (0,1,2) = per-allele effect coded by C-allele dosage. n = sample size.

**Table 5.**
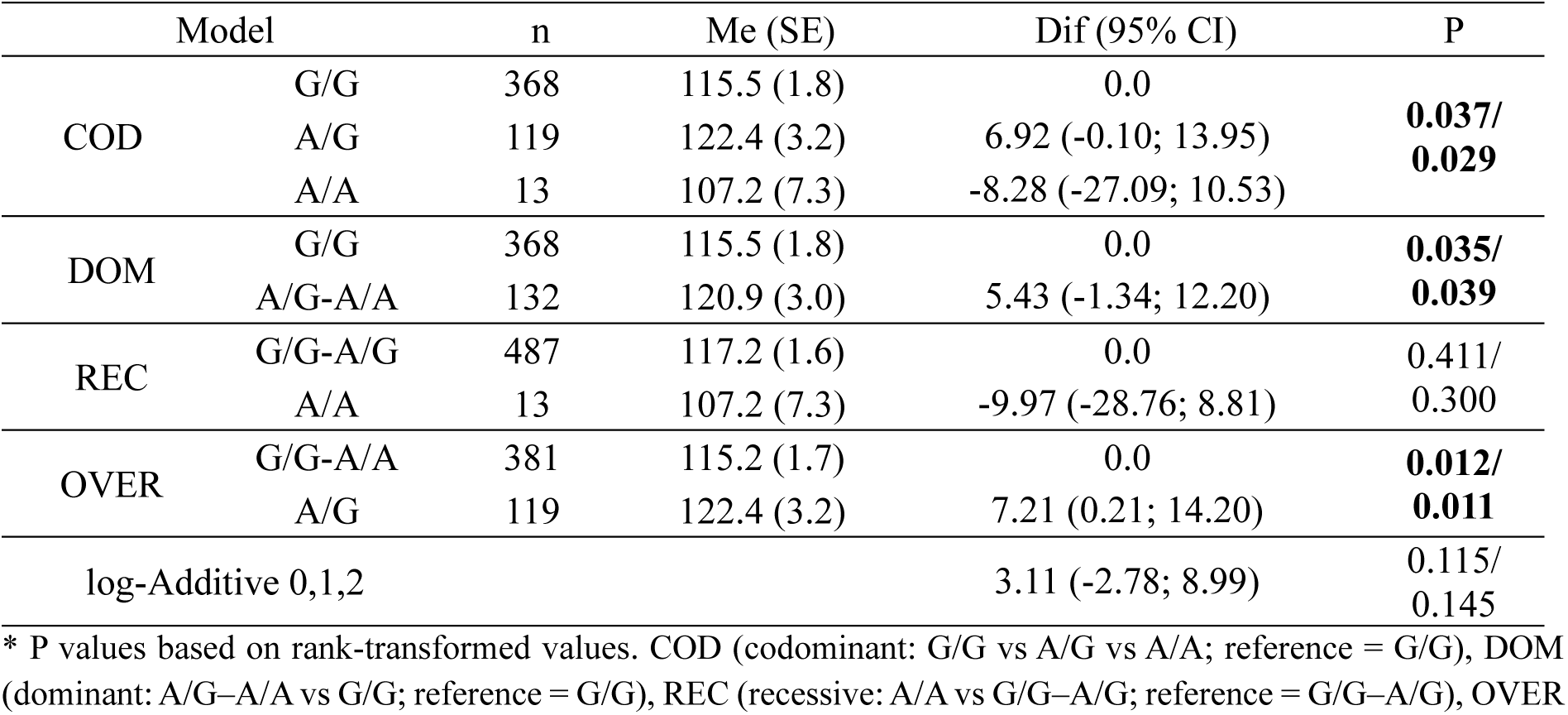
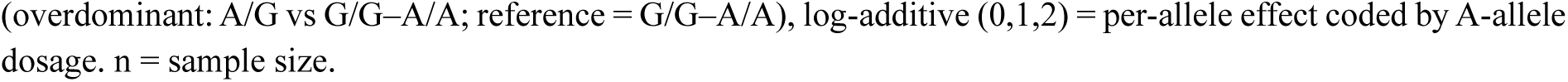
Association of *TNFα* rs1800629 with LDL cholesterol (mg·dL^-1^)

|  | Model | n | Me (SE) | Dif (95% CI) | P |
| --- | --- | --- | --- | --- | --- |
| COD | G/G | 368 | 115.5 (1.8) | 0.0 | **0.037/**<br>**0.029** |
|  | A/G | 119 | 122.4 (3.2) | 6.92 (-0.10; 13.95) |  |
|  | A/A | 13 | 107.2 (7.3) | -8.28 (-27.09; 10.53) |  |
| DOM | G/G | 368 | 115.5 (1.8) | 0.0 | **0.035/**<br>**0.039** |
|  | A/G–A/A | 132 | 120.9 (3.0) | 5.43 (-1.34; 12.20) |  |
| REC | G/G–A/G | 487 | 117.2 (1.6) | 0.0 | 0.411/<br>0.300 |
|  | A/A | 13 | 107.2 (7.3) | -9.97 (-28.76; 8.81) |  |
| OVER | G/G–A/A | 381 | 115.2 (1.7) | 0.0 | **0.012/**<br>**0.011** |
|  | A/G | 119 | 122.4 (3.2) | 7.21 (0.21; 14.20) |  |
| log-Additive 0,1,2 |  |  |  | 3.11 (-2.78; 8.99) | 0.115/<br>0.145 |

\* P values based on rank-transformed values. COD (codominant: G/G vs A/G vs A/A; reference = G/G), DOM (dominant: A/G–A/A vs G/G; reference = G/G), REC (recessive: A/A vs G/G–A/G; reference = G/G–A/G), OVER
(overdominant: A/G vs G/G–A/A; reference = G/G–A/A), log-additive (0,1,2) = per-allele effect coded by A-allele dosage. n = sample size.

**Table 6.** Association of *TNFα* rs1800629 with fitness status (LF vs HF)

| Model |  | LF N (%) | HF N (%) | OR (95% CI) | P |
| --- | --- | --- | --- | --- | --- |
| COD | G/G | 126 (66.3) | 243 (78.1) | 1.00 | 0.006/<br>0.090 |
|  | A/G | 60 (31.6) | 59 (19.0) | 0.51 (0.34; 0.78) |  |
|  | A/A | 4 (2.1) | 9 (2.9) | 1.17 (0.35; 3.86) |  |
| DOM | G/G | 126 (66.3) | 243 (78.1) | 1.00 | 0.004/<br>0.048 |
|  | A/G–A/A | 64 (33.7) | 68 (21.9) | 0.55 (0.37; 0.82) |  |
| REC | G/G–A/G | 186 (97.9) | 302 (97.1) | 1.00 | 0.585/<br>0.660 |
|  | A/A | 4 (2.1) | 9 (2.9) | 1.39 (0.42; 4.56) |  |
| OVER | G/G–A/A | 130 (68.4) | 252 (81.0) | 1.00 | 0.001/<br>0.030 |
|  | A/G | 60 (31.6) | 59 (19.0) | 0.51 (0.33; 0.77) |  |
| log-Additive 0,1,2 |  | 190 (37.9) | 311 (62.1) | 0.66 (0.46; 0.93) | 0.019/<br>0.106 |

\* P values based on rank-transformed values. COD (codominant: G/G vs A/G vs A/A; reference = G/G), DOM (dominant: A/G–A/A vs G/G; reference = G/G), REC (recessive: A/A vs G/G–A/G; reference = G/G–A/G), OVER (overdominant: A/G vs G/G–A/A; reference = G/G–A/A), log-additive (0,1,2) = per-allele effect coded by A-allele dosage.

### 3.2 Multi-locus analysis - Haplotype-based analysis IL6 and targeted gene-gene interactions

We estimated haplotypes from the three *IL6* SNPs and tested haplotype–phenotype association. In the *IL6* locus (rs1800795–rs1800796–rs1800797), six haplotypes were observed. The most frequent haplotype was G–G–G (frequency 0.487), followed by C–G–G (0.338), C–G–A (0.111), and G–C–G (0.063). Two haplotypes were rare (G–G–A, 0.0011; C–C–G, ∼0) and were excluded from haplotype-based association models. The G–G–G haplotype was used as the reference in regression analyses.

Among three prespecified phenotypes exhibiting a single-locus association with any of the *IL6* variants (i.e. iron, HCT), the *IL6* haplotype block showed a significant global association with iron (FDR-adjusted P (Q) = 0.032), whereas the association with haematocrit was not significant (Q = 0.084). For iron, follow-up haplotype-specific regression (reference G–G–G; rare haplotypes <1% excluded) C-G-G haplotype was associated with higher iron (β = 13.9, SE = 4.27, P= 0.0012; FDR-adjusted within-phenotype P = 0.002). Other common haplotypes (C-G-A and G-C-G) were not significant (FDR-adjusted within-phenotype P = 0.633). In the exploratory screening, we applied the *IL6* haplotype omnibus test (df = 4) across 43 continuous phenotypes, excluding haplotypes with frequency <1%. After Benjamini-Hochberg FDR correction across all exploratory phenotypes, no associations remained significant (minimum FDR-adjusted P= 0.976 for creatinine). Several traits showed nominal evidence (raw P < 0.05) - notably serum creatinine (P= 0.033) and platelets (PLT) (P= 0.045) - but none survived multiple-testing correction (thus no haplotype-specific models were fit for these parameter). Complete results of the exploratory haplotype-based analysis are presented in Table 8_SupplInfo.

Targeted gene-gene interaction analyses were pre-specified for the union of phenotypes showing single-locus signals (iron, HCT, LDL, VO2max mean). For each phenotype we added a block of *IL6*×*IL15* (and separately *IL6*×*TNFα*) interaction terms for the most frequent (>1% for continuous, and >10% for VO2max) non-reference *IL6* haplotypes and compared models with and without these terms using block likelihood-ratio tests. Interaction blocks (*IL6×IL15* and *IL6×TNFα*) were not significant (Table 9_SupplInfo).

To further potential non-additive and higher order interactions and higher-order interactions between loci beyond the predefined haplotype and pairwise interaction framework, we extended the analysis using a model-free approach (logic regression) to jointly evaluate SNPs from the *IL6*, *IL15*, and *TNF* loci in relation to VO2max (Figure 1).

**Figure 1.**
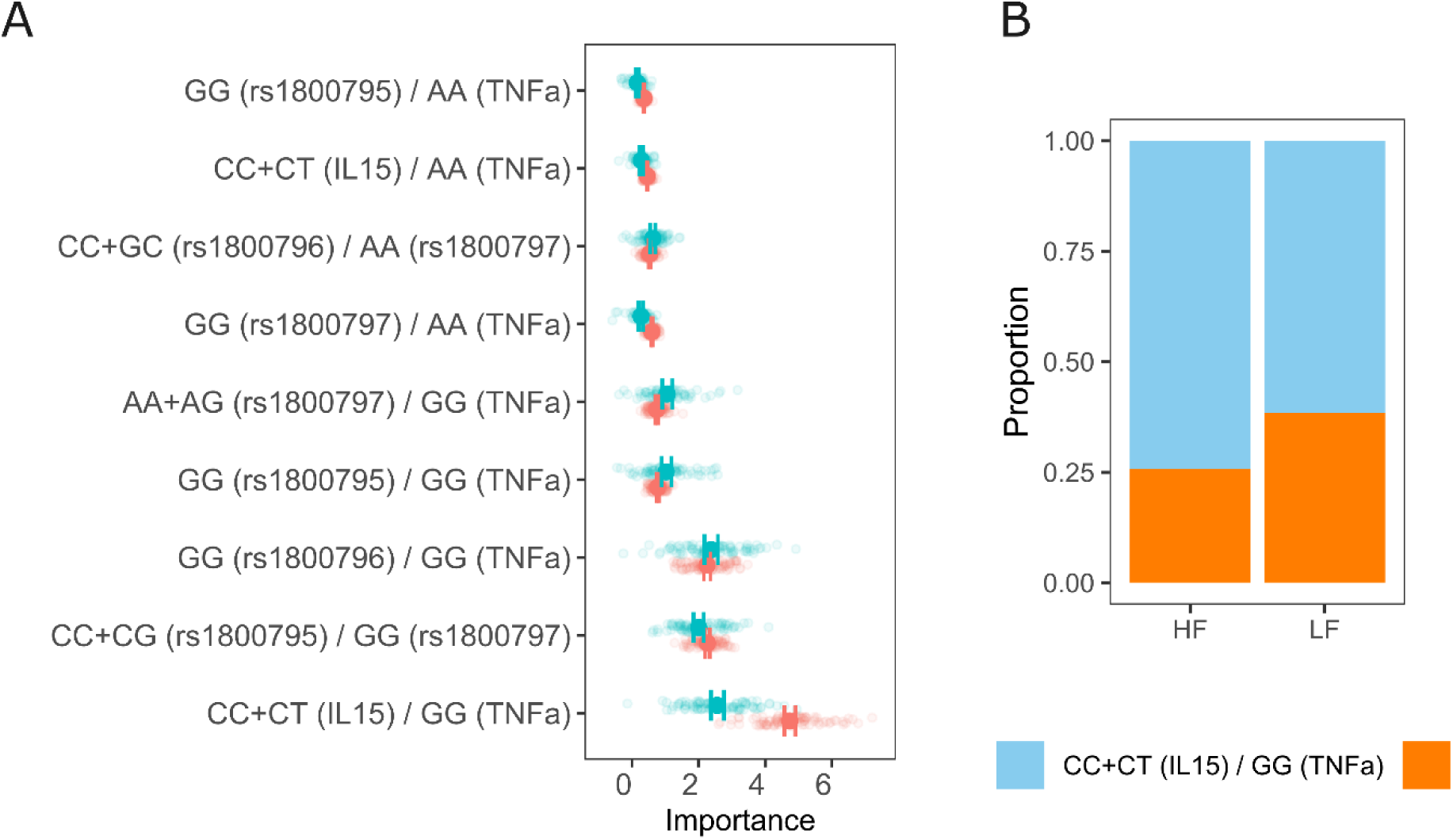
Data-driven multi-locus analysis (*IL6*, *IL15*, *TNF-a*) association analysis with VO2max status

Multi-locus logic regression analysis identified the genotype combination *IL15* CC+CT and *TNF*-α GG as the most influential, with a median importance of 4.74 (95% CI: 4.57-4.91) in real data, compared to 2.55 (95% CI: 2.36-2.75) in resampled data (Figure 1A). This combination was significantly more frequent in the HF group than in the LF group (Chi-square p = 0.004, Figure 1B).

## 4. DISCUSSION

To address the question of whether genetic variation, particularly in cytokine regulation, interacts with inflammatory markers, body composition, basic biochemical parameters, and variables of aerobic and anaerobic performance, the individual and combined roles of five SNPs in *IL6* (rs1800795, rs1800796, rs1800797), *IL15* (rs1589241), and *TNF-α* (rs1800629) genes were investigated in the present highly complex and multidimensional study. A major strength of this study is the large cohort of 1000 healthy adults of Caucasian origin who underwent comprehensive physiological, biochemical, metagenomic, and genomic assessments. Another important strength lies in the innovative classification strategy, instead of comparing athletes with non-athletes, participants were grouped according to objectively measured aerobic capacity (VO2max) based on standardized norms. When combined with the analysis of five single-locus polymorphisms, haplotypes, and gene-gene interactions, these features constitute a unique contribution of this study to the field.

When comparing basic physiological and biochemical variabilities between the participants with very good or outstanding VO2max and participants with satisfactory, poor, or very poor VO2max, clear differences were observed. Participants who had higher VO2max values exhibited healthier lipid profiles and lower white blood cell counts, suggesting reduced baseline inflammation. They also presented with lower body mass and waist circumference, reflecting more favourable metabolic health. Interestingly, although their red blood cell and haemoglobin levels were significantly lower than those of the control group, the HF group demonstrated higher mean corpuscular volume (MCV) and mean corpuscular haemoglobin (MCH), indicating fewer but larger erythrocytes, which may represent a haematological adaptation. Numerous studies have confirmed significant training-induced changes in body mass and composition, as well as in physiological and biochemical parameters. These typically include reductions in body weight, fat mass, and skinfold thickness; increases in VO2max and cardiorespiratory efficiency; favourable alterations in haematological indices (HCT, MCV, MCHC, WBC, neutrophils, monocytes, MPV); and improvements in glucose homeostasis and lipid profile [21–23]. In addition, regular exercise is associated with reduced resting heart rate and blood pressure, improved endothelial function, increased muscle capillarization and mitochondrial density, as well as favourable modulation of inflammatory and oxidative stress markers (e.g., CRP, IL6, TNF-α) [24–26]. These findings raise the question of whether genetic variation, particularly in cytokine regulation, may help explain these patterns of post-training body changes. Our data are consistent with this inflammatory–regenerative axis: the High Fitness group displayed lower WBC and a markedly more favourable HDL/LDL/TG profile alongside superior aerobic and anaerobic metrics, which provides a phenotypic backdrop for testing cytokine-pathway genetics.

To examine the influence of these genetic variables, firstly three functional polymorphisms (rs1800795, rs1800796, rs1800797) located in the promoter region of the *IL6* gene were investigated. All of these variants are suggested to influence IL6 expression [27] and consequently modulate post-exercise inflammation and recovery [27,28]. For example, the C allele of rs1800795 (–174G/C) has been associated with significantly reduced IL6 serum concentrations in response to long-term exercise training programs [29]. Although, the molecular analysis conducted in this study did not reveal significant associations between the individual and combined *IL6* polymorphisms and VO2max, genotype-specific differences in blood parameters were observed across the cohort. Individuals with the CC genotype of rs1800795 exhibited slightly higher haematocrit and iron levels. These differences suggest that the CC genotype may support more efficient recovery and metabolic stability, even if its direct effect on aerobic performance is limited. In contrast, the other two *IL6* promoter polymorphisms showed no significant associations with the blood-based markers assessed. However, the *IL6* haplotype block showed a significant global association with iron, whereas the association with haematocrit was not significant. Specifically, C-G-G (rs1800795– rs1800796–rs1800797) haplotype was associated with higher iron, while other common haplotypes (C-G-A and G-C-G) were not significant. These genetic findings complement evidence from non-genetic markers, such as cfDNA responses to maximal exercise [12] and adipokine dynamics modulated by fat mass and exercise intensity [13], both highlighting the role of inflammatory and metabolic axes in shaping recovery. Consistent with our findings, other authors have confirmed the association of the rs1800795 polymorphism with various biochemical blood parameters. Specifically, the CC genotype has been linked to lower circulating IL6 levels and reduced systemic inflammation [30], as well as to differences in glucose metabolism and lipid profile in several cohorts [23,31]. In addition, recent evidence indicates that IL6 signalling contributes to iron metabolism through the IL6/JAK1/STAT3 pathway, which upregulates divalent metal transporter 1 (*DMT1*) expression via increased hypoxia-inducible factor-1 α (HIF-1α) activity [32]. This mechanism may also be relevant in athletes, as exercise-induced fluctuations in IL6 could transiently alter iron homeostasis and erythropoiesis, thereby influencing haematological adaptations to training. In our single-locus models, rs1800795 showed nominal associations with iron (dominant model Δ≈+15 µg·dL^-1^ for C-allele carriers) and haematocrit, but not with VO2max. These signals require confirmation in covariate-adjusted and multiplicity-controlled analyses.

Secondly, the rs1800629 polymorphism located in the proximal promoter region of *TNF-α* was examined. *TNF-α* has a dual role in skeletal muscle: brief signalling helps initiate myogenesis and tissue repair, whereas sustained or excessive TNF-α drives NF-κB-ubiquitin-proteasome catabolism, promotes chronic inflammation, and delays recovery-ultimately impairing performance [33,34]. In the present single-locus analysis, individuals carrying the AG or AA genotypes were significantly less likely to be classified as participants with higher VO2max values. This finding supports the hypothesis that the A allele may represent a disadvantageous factor for aerobic fitness, likely due to its association with elevated *TNF-α* expression, which may delay recovery or hinder training adaptations. This aligns with functional evidence that the rs1800629 A allele increases TNF-α transcriptional activity, particularly under conditions of stress or immune activation [35]. Elevated TNF-α level can impede endurance adaptations by inducing skeletal-muscle insulin resistance, suppressing protein synthesis, and perturbing myogenesis, thereby delaying recovery [36–38] . Consistent with this, rs1800629 modifies the anti-inflammatory benefits of physical activity such as smaller CRP reductions in A allele carriers [39] and has been linked to poorer physical-function performance in population cohorts [40], while also modulating training-induced biochemical adaptations [41], together supporting the hypothesis that the A allele may be disadvantageous for aerobic fitness. Beyond aerobic capacity, we observed a suggestive trend for longer Wingate time to peak power in AA versus AG/GG (p=0.052) (Table 7_SupplInfo), directionally consistent with prior data that elevated TNF-α can impair excitation–contraction coupling and recovery [42–44], thereby slowing neuromuscular activation and diminishing acute power output. This points to a TNF-linked lipid-neuromuscular signature that fits a pro-inflammatory environment. Together, these findings suggest that cytokine-related polymorphisms influence distinct components of the exercise response - *IL6* variants linked primarily to iron handling and haematological adaptation, and *TNF*-α variants to inflammatory tone and performance recovery. Similarly, recent omics-based research shows that gut microbiome profiles differentiate responses to aerobic versus anaerobic exercise, along with circulating biomarkers [45]. These findings support the idea that the inflammatory-genetic-microbial axis is a key factor in individual adaptation.

The next polymorphism examined was rs1589241, located in a regulatory intronic region of the *IL15* gene, which encodes an important exercise-induced cytokine involved in immune regulation, fat metabolism, mitochondrial biogenesis, and skeletal muscle growth [46–48]. Our single-locus analysis revealed no significant associations between the *IL15* genotypes and VO2max or biochemical variabilities. However, notable findings emerged when gene–gene interactions were modelled using a multi-locus logistic regression. Specifically, individuals carrying the *IL15* CC or CT genotypes together with the *TNF-α* GG genotype were significantly more likely to be classified as participants with higher VO2max. This suggests a potential synergistic interaction in which a favourable *IL15* background acts in concert with a low-producer TNF-α profile to facilitate recovery and training-induced aerobic changes. Biological plausibility for *IL15* in endurance adaptation is strong, in detail, this training upregulates muscular *IL15* in humans, which promotes myogenesis and mitigating inflammation-induced atrophy [46,48,49]. Although the interaction between these genes and VO2max has not been studied so far, the relationship between these cytokines is well described. IL15 and TNF-α exert opposing influences on skeletal muscle: TNF-α promotes catabolic signalling and impairs contractile function, whereas IL15 is myokine with pro-myogenic and anti-atrophic actions. Notably, human myotubes secrete IL15 in response to TNF-α, consistent with a compensatory, anti-inflammatory feedback loop [44,46]. IL15 attenuates TNF-α–driven proteolysis and apoptosis in muscle and can inhibit downstream catabolic TNF-α pathways (e.g., NF-κB/UPS), thereby preserving muscle mass and function [48,50]. In athletes, endurance training increases skeletal-muscle IL15 protein content, positioning IL15 as a training-responsive mediator of adaptation [49]. Conversely, dampening TNF-α signalling facilitates recovery after injury, underscoring the performance cost of a pro-inflammatory environment of TNF-α) [50]. Taken together, these data support a synergistic model in which *IL15* variation that favours robust IL15 signalling may be most beneficial when combined with a low-producer TNF-α background (e.g., rs1800629 GG), helping to reduce inflammation, speed recovery, and ultimately enhance endurance adaptations [48,49,50]. These interaction findings are preliminary and warrant replication with full covariate adjustment and correction for multiple testing.

Collectively, these findings highlight the potential value of integrating genetic markers with physiological and biochemical profiling when investigating individual variability in exercise phenotype. While current effect sizes are modest, such integrative approaches may contribute to more refined strategies for training personalization, recovery monitoring, and targeted interventions in both athletic and clinical settings.

## 5. LIMITATIONS

There are several limitations of this study that should be acknowledged. First, the cross-sectional design precludes drawing causal inferences about training responses or long-term adaptations. Second, while testing was standardized, residual confounding factors such as diet, recent illness, or circadian variation cannot be fully excluded. Third, this was a single-country cohort of adults of European (Polish) ancestry, which limits generalizability to other ancestries, regions, and sport populations; replication in more diverse, multi-centre cohorts is needed. Fourth, the candidate-gene approach focused on biologically plausible *loci*, but it does not capture genome-wide variation. Thus, the contribution of other variants remains unexplored. Finally, rare genotype groups (e.g., *TNF-α* rs1800629 AA) limited the power of recessive models, and deviations from the HWE at some *loci* require cautious interpretation. Prospective and longitudinal studies are required to validate these findings.

## 6. CONCLUSIONS

Taken together, the results of this study provide a multilevel picture of how cytokine-related genes may contribute to variability in both aerobic and anaerobic performance traits. In a large cohort of 1,000 healthy adults, *IL6* rs1800795 showed associations with haematological traits linked to recovery and iron handling (higher haematocrit and serum iron in C-allele carriers), whereas the *TNF-α* rs1800629 A allele appeared to be disadvantageous for both aerobic and probably anaerobic performance (longer time to reach peak power in AA homozygotes). Importantly, multi-locus pattern was also observed: the *IL15* CC/CT × *TNF-α* GG combination was associated with greater odds of higher aerobic capacity. These results highlight the potential value of considering gene-gene combinations alongside single-variant models and integrating genetic markers with physiological profiling. Because several signals were modest or exploratory, and the study is cross-sectional in a European cohort, replication with covariate adjustment and multiplicity control in independent, longitudinal samples is warranted to define effect sizes and generalisability.

## Funding information

This research was conducted as a part of the commissioned research project ZUB 1/2020 - "1000 Genomes: Genetic Basis of Physical Activity, Sport Level, and Human Well-Being" and co-financed from the state budget under the program of the Minister of Education and Science, "Science for Society II", project no. NdS-II/SP/0503/2024/01. The amount of the grant was 1 million PLN and the total value of the project was 1 million PLN.

A.V.S was supported by funding by the South African Medical Research Council through its Division of Research Capacity Development under the Mid-Career Scientist Programme, from funding received from the South African National Treasury. The content hereof is the sole responsibility of the authors and does not necessarily represent the official view of the SAMRC or the funders.

## Ethics approval

The study complied with the Declaration of Helsinki and was approved by the Bioethics Committee at the District Medical Chamber in Gdańsk (KB-16/20).

## Conflicts of interest

The authors declare no conflicts of interest.

## Data availability statement

All data produced in the present study are available upon reasonable request to the authors

## Authors Contribution

Conceptualization, K.H-L; Data curation, K.H-L, P.A., A.L-D; Formal analysis, K.H-L, P.A; Funding acquisition: K.H-L.; Investigation, K.H-L., B.B, M.M-S., P.A.; Methodology, K.H-L, B.B, M.M.-S., P.A.; Project administration, K.H-L, M.M-S.; Resources: K.H-L., P.A.; Software: K.H-L., A.V.S.; Supervision, K.H-L.; Validation, K.H-L., B.B, M.M-S., P.P.; Visualization: K.H-L., B.B.; Writing – original draft, K.H-L., A.L-D.; Writing – review & editing, K.H-L., A.V.S, B.B., P.P, A.L-D.

## Acknowledgments

The authors thank all the volunteers who enthusiastically participated in this research project.

## Notes

### Competing Interest Statement

The authors have declared no competing interest.

