## Supplemental Tables for "Cytokine Gene Variants Are Associated with the Inflammatory and Metabolic Profile of Human Exercise Performance"

Table 1_SupplInfo. Aerobic capacity standards based on $VO_2$max levels adapted from Shvarz and Reibold, 1990.

| Men | | | | | | | |
|---|---|---|---|---|---|---|---|
| age | very poor | poor | satisfactory | medium | good | very good | outstanding |
| 20-24 | <32 | 32-37 | 38-43 | 44-50 | 51-56 | 57-62 | >62 |
| 25-29 | <31 | 31-35 | 36-42 | 43-48 | 49-53 | 54-59 | >59 |
| 30-34 | <29 | 29-34 | 35-40 | 41-45 | 46-51 | 52-56 | >56 |
| 35-39 | <28 | 28-32 | 33-38 | 39-43 | 44-48 | 49-54 | >54 |
| 40-44 | <26 | 26-31 | 32-35 | 36-41 | 42-46 | 47-51 | >51 |
| 45-49 | <25 | 25-29 | 30-34 | 35-39 | 40-43 | 44-48 | >48 |
| 50-54 | <24 | 24-27 | 28-32 | 33-36 | 37-41 | 42-46 | >46 |
| 55-59 | <22 | 22-26 | 27-30 | 31-34 | 35-39 | 40-43 | >43 |
| 60-65 | <21 | 31-24 | 25-28 | 29-32 | 33-36 | 37-40 | >40 |
| Woman | | | | | | | |
| age | very poor | poor | satisfactory | medium | good | very good | outstanding |
| 20-24 | <27 | 27-31 | 32-36 | 37-41 | 42-46 | 47-51 | >51 |
| 25-29 | <26 | 26-30 | 31-35 | 36-40 | 41-44 | 45-49 | >49 |
| 30-34 | <25 | 25-29 | 30-33 | 34-37 | 38-42 | 43-46 | >46 |
| 35-39 | <24 | 24-27 | 28-31 | 32-35 | 36-40 | 41-44 | >44 |
| 40-44 | <22 | 22-25 | 26-29 | 30-33 | 34-37 | 38-41 | >41 |
| 45-49 | <21 | 21-23 | 24-27 | 28-31 | 32-35 | 36-38 | >38 |
| 50-54 | <19 | 19-22 | 23-25 | 26-29 | 30-32 | 33-36 | >36 |
| 55-59 | <18 | 18-20 | 21-23 | 24-27 | 28-30 | 31-33 | >33 |
| 60-65 | <16 | 16-18 | 19-21 | 22-24 | 25-27 | 28-30 | >30 |

Sex- and age-specific $VO_2$max cut-offs ($mL \cdot kg^{-1} \cdot min^{-1}$) for men and women aged 20–65 years, categorized as: very poor, poor, satisfactory, medium, good, very good, outstanding.

Table 2_SupplInfo. Hardy-Weinberg equilibrium

| SNP | Combined (LF+HF) | LF | HF |
|---|---|---|---|
| IL15 (rs1589241) | 0.022 | 0.390 | 0.745 |
| IL6 (rs1800795) | 0.845 | 0.884 | 0.819 |
| IL6 (rs1800796) | 0.578 | 1.0 | 1.0 |
| IL6 (rs1800797) | 0.00018 | 0.138 | 0.398 |
| TNFα (rs1800629) | 0.804 | 0.457 | **0.034** |

P values obtained from an exact SNP test of Hardy-Weinberg Equilibrium, computed using the tableHWE function in the R snpassoc package. Values <0.05 indicate deviation from HWE. LF, Low Fitness; HF, High Fitness (defined from age- and sex-specific $VO_2$max categories).

Table 3_SupplInfo. *IL6* 1800795 (N=501)

| **Characteristic** | **CC** N = 100[1] | **CG** N = 250[1] | **GG** N = 151[1] | **P**[2] |
|---|---|---|---|---|
| Glucose ($mg \cdot dL^{-1}$) | 93 (8) | 96 (10) | 95 (9) | 0.051 |
| Creatinine ($mg \cdot dL^{-1}$) | 0.90 (0.14) (N=98) | 0.91 (0.14) | 0.88 (0.16) | 0.147 |

| | | | | | |
|---|---|---|---|---|---|
| Iron (μg·dL⁻¹) | | 116 (45) (N=99) | 122 (49) | 105 (42) (N=150) | **0.004** |
| WBC ($10^9$·$L^{-1}$) | | 5.75 (1.42) | 5.60 (1.50) | 5.66 (1.77) | 0.493 |
| RBC ($10^{12}$·$L^{-1}$) | | 4.91 (0.47) | 4.94 (0.41) | 4.84 (0.42) | 0.051 |
| HGB (g·$dL^{-1}$) | | 14.70 (1.36) | 14.85 (1.16) | 14.53 (1.27) | 0.072 |
| HCT (%) | | 43.69 (3.50) | 43.96 (2.97) | 43.04 (3.17) | **0.033** |
| MCV (fL) | | 89.3 (4.0) | 89.2 (3.9) | 89.1 (3.9) | 0.995 |
| MCH (pg) | | 30.01 (1.56) | 30.11 (1.44) | 30.04 (1.45) | 0.806 |
| MCHC (g·$dL^{-1}$) | | 33.63 (0.98) | 33.78 (0.88) | 33.72 (0.90) | 0.548 |
| PLT ($10^9$·$L^{-1}$) | | 236 (51) | 232 (46) | 237 (51) (N=150) | 0.552 |
| TC (mg·$dL^{-1}$) | | 192 (33) | 189 (36) | 190 (38) (N=150) | 0.686 |
| HDL (mg·$dL^{-1}$) | | 69 (18) (N=99) | 67 (18) | 66 (18) | 0.577 |
| LDL (mg·$dL^{-1}$) | | 121 (30) (N=99) | 116 (34) | 117 (36) | 0.348 |
| TG (mg·$dL^{-1}$) | | 92 (58) (N=99) | 103 (63) | 101 (60) (N=150) | 0.217 |
| Cortisol (μg·$dL^{-1}$) | | 16.1 (5.1) (N=99) | 16.1 (5.6) | 15.5 (5.2) | 0.629 |
| $VO_2$max (mL/min/kg) | LF | 39 / 100 (39%) | 93 / 250 (37%) | 58 / 151 (38%) | 0.942 |
| | HF | 61 / 100 (61%) | 157 / 250 (63%) | 93 / 151 (62%) | |
| Body Mass (kg) | | 76 (14) | 78 (15) | 76 (15) | 0.272 |
| Waist Circumference (cm) | | 82 (11) | 85 (12) | 83 (11) | 0.249 |
| Hip Circumference (cm) | | 98 (7) | 99 (8) | 98 (7) | 0.099 |
| BMI (kg·$m^{-2}$) | | 24.2 (3.5) | 24.8 (3.9) | 24.4 (3.7) | 0.321 |
| HR (beats·$min^{-1}$) | | 69 (10) | 70 (13) (N=249) | 70 (14) | 0.925 |
| Sex | Females | 31 / 100 (31%) | 74 / 250 (30%) | 57 / 151 (38%) | 0.228 |
| | Males | 69 / 100 (69%) | 176 / 250 (70%) | 94 / 151 (62%) | |
| Age (years) | | 36 (7) | 36 (8) | 35 (8) | 0.079 |
| Stage of Test Termination | | 5.67 (1.73) | 5.63 (1.93) | 5.59 (1.85) | 0.925 |
| Time of Test Termination (min) | | 15.0 (3.5) | 14.8 (3.9) | 14.7 (3.5) | 0.795 |
| HRmax (beats·$min^{-1}$) | | 179 (12) | 179 (13) | 181 (11) | 0.175 |
| VO2max(mL/min/kg) | | 45 (11) | 46 (13) | 45 (12) | 0.817 |
| VO2max(L/min) | | 3.36 (0.94) | 3.47 (0.95) | 3.39 (0.89) | 0.441 |
| VEmax (L/min) | | 128 (38) | 132 (37) | 127 (37) | 0.505 |
| RER_max | | 1.19 (0.07) | 1.19 (0.07) | 1.19 (0.08) | 0.971 |
| Workload at Termination (W/kg) | | 3.83 (0.87) | 3.82 (0.97) | 3.80 (0.93) | 0.927 |

| | | | | |
|---|---|---|---|---|
| End of Test | 18.0 (3.5) | 17.8 (3.9) | 17.7 (3.5) | 0.795 |
| Fatigue Index | 0.02 (0.14) (N=95) | 0.02 (0.11) (N=241) | 0.03 (0.12) (N=145) | 0.413 |
| Average Power (W) | 545 (141) | 563 (136) | 542 (144) | 0.350 |
| Relative Average Power (per Body Mass) (W·kg⁻¹) | 7.18 (1.10) | 7.24 (1.28) | 7.14 (1.25) | 0.682 |
| Maximum Power (W) | 687 (191) | 715 (176) | 686 (197) | 0.326 |
| Relative Maximum Power (per Body Mass) (W·kg⁻¹) | 9.00 (1.39) | 9.14 (1.46) | 8.98 (1.58) | 0.656 |
| Time to Reach Pmax (s) | 6.44 (2.00) | 6.61 (2.58) | 6.71 (3.09) | 0.879 |
| Maximum Rotational Speed (rev·s⁻¹) | 2.05 (0.29) | 2.08 (0.35) | 2.04 (0.36) | 0.705 |
| Strength-Speed Index | 86 (39) | 88 (36) | 84 (37) | 0.487 |

WBC, white blood cell count ($10^9$·$L^{-1}$); RBC, red blood cell count ($10^{12}$·$L^{-1}$); HGB, haemoglobin (g·$dL^{-1}$); HCT, haematocrit (%); MCV, mean corpuscular volume (fL); MCH, mean corpuscular haemoglobin (pg); MCHC, mean corpuscular haemoglobin concentration (g·$dL^{-1}$); PLT, platelets ($10^9$·$L^{-1}$); TC, total cholesterol (mg·$dL^{-1}$); HDL, high-density lipoprotein cholesterol (mg·$dL^{-1}$); LDL, low-density lipoprotein cholesterol (mg·$dL^{-1}$); TG, triglycerides (mg·$dL^{-1}$); HR, heart rate (beats·$min^{-1}$); HRmax, maximal heart rate (beats·$min^{-1}$); $VO_2max$, maximal oxygen uptake reported as mL·$kg^{-1}$·$min^{-1}$ (mass-normalized) and L·$min^{-1}$ (absolute); VEmax, maximal minute ventilation (L·$min^{-1}$); RER_max, maximal respiratory exchange ratio; Workload at Termination, final incremental load (W·$kg^{-1}$); Stage/Time of Test Termination, stage number and elapsed time (min) at cessation of the incremental test; End of Test, rating of perceived exertion (Borg 6–20); Fatigue Index, Wingate-derived fatigue metric; Average Power (W) and Relative Average Power (W·$kg^{-1}$); Maximum Power (W) and Relative Maximum Power (W·$kg^{-1}$); Time to Reach Pmax, time to peak power (s); Maximum Rotational Speed (rev·$s^{-1}$).

Table 4_SupplInfo. *IL6* 1800796 (N=501)

| **Characteristic** | **CC + GC** N = 62[1] | **GG** N = 439[1] | **P**[2] |
|---|---|---|---|
| Glucose (mg·dL⁻¹) | 96 (12) | 95 (9) | 0.806 |
| Creatinine (mg·dL⁻¹) | 0.91 (0.13) | 0.90 (0.15) (N=437) | 0.423 |
| Iron (µg·dL⁻¹) | 117 (53) | 116 (46) (N=437) | 0.697 |
| WBC ($10^9$·L⁻¹) | 5.54 (1.71) | 5.66 (1.55) | 0.504 |
| RBC ($10^{12}$·L⁻¹) | 4.88 (0.35) | 4.91 (0.43) | 0.548 |
| HGB (g·dL⁻¹) | 14.69 (1.11) | 14.73 (1.26) | 0.591 |
| HCT (%) | 43.50 (2.77) | 43.64 (3.22) | 0.592 |
| MCV (fL) | 89.2 (4.2) | 89.2 (3.9) | 0.617 |
| MCH (pg) | 30.11 (1.54) | 30.06 (1.46) | 0.640 |
| MCHC (g·dL⁻¹) | 33.75 (0.96) | 33.73 (0.90) | 0.891 |
| PLT ($10^9$·L⁻¹) | 226 (47) | 235 (49) (N=438) | 0.101 |
| TC (mg·dL⁻¹) | 192 (37) | 190 (36) (N=437) | 0.526 |

| | | | | |
|---|---|---|---|---|
| HDL (mg·dL⁻¹) | | 67 (19) | 67 (18) (N=438) | 0.838 |
| LDL (mg·dL⁻¹) | | 118 (34) | 117 (34) (N=438) | 0.616 |
| TG (mg·dL⁻¹) | | 100 (65) | 100 (61) (N=437) | 0.440 |
| Cortisol (µg·dL⁻¹) | | 15.6 (4.4) | 16.0 (5.5) (N=438) | 0.952 |
| $VO_2$max (mL/min/kg) | LF | 23 / 62 (37%) | 167 / 439 (38%) | 0.997 |
| | HF | 39 / 62 (63%) | 272 / 439 (62%) | |
| Body Mass (kg) | | 75 (14) | 77 (15) | 0.512 |
| Waist Circumference (cm) | | 83 (10) | 84 (12) | 0.818 |
| Hip Circumference (cm) | | 98 (7) | 99 (7) | 0.847 |
| BMI (kg·m⁻²) | | 24.1 (3.3) | 24.6 (3.8) | 0.415 |
| HR (beats·min⁻¹) | | 69 (15) | 70 (12) (N=438) | 0.517 |
| Sex | Females | 19 / 62 (31%) | 143 / 439 (33%) | 0.874 |
| | Males | 43 / 62 (69%) | 296 / 439 (67%) | |
| Age (years) | | 36 (8) | 36 (8) | 0.873 |
| Stage of Test Termination | | 5.63 (1.63) | 5.63 (1.90) | 0.952 |
| Time of Test Termination (min) | | 14.8 (3.3) | 14.8 (3.8) | 0.947 |
| HRmax (beats·min⁻¹) | | 181 (12) | 179 (13) | 0.524 |
| VO2max(mL/min/kg) | | 45 (12) | 45 (12) | 0.797 |
| VO2max(L/min) | | 3.41 (0.96) | 3.43 (0.92) | 0.794 |
| VEmax (L/min) | | 134 (36) | 129 (37) | 0.315 |
| RER_max | | 1.20 (0.07) | 1.19 (0.07) | 0.594 |
| Workload at Termination (W/kg) | | 3.82 (0.82) | 3.82 (0.95) | 0.973 |
| End of Test | | 17.8 (3.3) | 17.8 (3.8) | 0.947 |
| Fatigue Index | | 0.01 (0.11) (N=61) | 0.02 (0.12) (N=420) | 0.636 |
| Average Power (W) | | 553 (137) | 553 (140) | 0.885 |
| Relative Average Power (per Body Mass) (W·kg⁻¹) | | 7.33 (1.09) | 7.18 (1.26) | 0.356 |
| Maximum Power (W) | | 695 (179) | 701 (187) | 0.948 |
| Relative Maximum Power (per Body Mass) (W·kg⁻¹) | | 9.18 (1.33) | 9.05 (1.51) | 0.595 |
| Time to Reach Pmax (s) | | 6.50 (3.28) | 6.62 (2.54) | 0.398 |

| | | | |
|---|---|---|---|
| Maximum Rotational Speed ($rev \cdot s^{-1}$) | 2.08 (0.30) | 2.06 (0.35) | 0.592 |
| Strength-Speed Index | 85 (33) | 87 (37) | 0.991 |

WBC, white blood cell count ($10^9 \cdot L^{-1}$); RBC, red blood cell count ($10^{12} \cdot L^{-1}$); HGB, haemoglobin ($g \cdot dL^{-1}$); HCT, haematocrit (%); MCV, mean corpuscular volume (fL); MCH, mean corpuscular haemoglobin (pg); MCHC, mean corpuscular haemoglobin concentration ($g \cdot dL^{-1}$); PLT, platelets ($10^9 \cdot L^{-1}$); TC, total cholesterol ($mg \cdot dL^{-1}$); HDL, high-density lipoprotein cholesterol ($mg \cdot dL^{-1}$); LDL, low-density lipoprotein cholesterol ($mg \cdot dL^{-1}$); TG, triglycerides ($mg \cdot dL^{-1}$); HR, heart rate ($beats \cdot min^{-1}$); HRmax, maximal heart rate ($beats \cdot min^{-1}$); $VO_2max$, maximal oxygen uptake reported as $mL \cdot kg^{-1} \cdot min^{-1}$ (mass-normalized) and $L \cdot min^{-1}$ (absolute); VEmax, maximal minute ventilation ($L \cdot min^{-1}$); RER_max, maximal respiratory exchange ratio; Workload at Termination, final incremental load ($W \cdot kg^{-1}$); Stage/Time of Test Termination, stage number and elapsed time (min) at cessation of the incremental test; End of Test, rating of perceived exertion (Borg 6–20); Fatigue Index, Wingate-derived fatigue metric; Average Power (W) and Relative Average Power ($W \cdot kg^{-1}$); Maximum Power (W) and Relative Maximum Power ($W \cdot kg^{-1}$); Time to Reach Pmax, time to peak power (s); Maximum Rotational Speed ($rev \cdot s^{-1}$).

Table 5_SupplInfo. *IL6* 1800797 (N=501)

| **Characteristic** | | **AA_AG** N = 110[1] | **GG** N = 391[1] | **P**[2] |
|---|---|---|---|---|
| Glucose ($mg \cdot dL^{-1}$) | | 94 (10) | 96 (9) | 0.091 |
| Creatinine ($mg \cdot dL^{-1}$) | | 0.89 (0.14) (N=108) | 0.90 (0.15) | 0.553 |
| Iron ($\mu g \cdot dL^{-1}$) | | 116 (45) (N=109) | 116 (47) (N=390) | 0.813 |
| WBC ($10^9 \cdot L^{-1}$) | | 5.79 (1.43) | 5.60 (1.61) | 0.115 |
| RBC ($10^{12} \cdot L^{-1}$) | | 4.90 (0.47) | 4.90 (0.41) | 0.934 |
| HGB ($g \cdot dL^{-1}$) | | 14.70 (1.36) | 14.73 (1.20) | 0.899 |
| HCT (%) | | 43.66 (3.47) | 43.61 (3.07) | 0.695 |
| MCV (fL) | | 89.3 (3.9) | 89.1 (3.9) | 0.790 |
| MCH (pg) | | 30.05 (1.54) | 30.08 (1.45) | 0.787 |
| MCHC ($g \cdot dL^{-1}$) | | 33.64 (0.98) | 33.76 (0.89) | 0.395 |
| PLT ($10^9 \cdot L^{-1}$) | | 238 (51) | 233 (48) (N=390) | 0.315 |
| TC ($mg \cdot dL^{-1}$) | | 193 (31) (N=109) | 189 (37) (N=390) | 0.159 |
| HDL ($mg \cdot dL^{-1}$) | | 70 (18) (N=109) | 67 (18) | 0.068 |
| LDL ($mg \cdot dL^{-1}$) | | 120 (30) (N=109) | 116 (35) | 0.131 |
| TG ($mg \cdot dL^{-1}$) | | 95 (58) (N=109) | 102 (62) (N=390) | 0.312 |
| Cortisol ($\mu g \cdot dL^{-1}$) | | 16.2 (5.2) (N=109) | 15.8 (5.4) | 0.440 |
| $VO_2max$ (mL/min/kg) | LF | 43 / 110 (39%) | 147 / 391 (38%) | 0.862 |
| | HF | 67 / 110 (61%) | 244 / 391 (62%) | |
| Body Mass (kg) | | 76 (15) | 77 (15) | 0.802 |
| Waist Circumference (cm) | | 83 (13) | 84 (12) | 0.365 |

| | | | |
|---|---|---|---|
| Hip Circumference (cm) | 98 (8) | 98 (7) | 0.830 |
| BMI (kg·m⁻²) | 24.5 (4.0) | 24.6 (3.7) | 0.618 |
| HR (beats·min⁻¹) | 69 (10) | 70 (13) (N=390) | 0.904 |
| Sex: Females | 38 / 110 (35%) | 124 / 391 (32%) | 0.656 |
| Sex: Males | 72 / 110 (65%) | 267 / 391 (68%) | |
| Age (years) | 36 (7) | 36 (8) | 0.490 |
| Stage of Test Termination | 5.59 (1.79) | 5.64 (1.89) | 0.776 |
| Time of Test Termination (min) | 14.8 (3.6) | 14.8 (3.7) | 0.969 |
| HRmax (beats·min⁻¹) | 178 (13) | 180 (13) | 0.120 |
| VO2max(mL/min/kg) | 44 (11) | 46 (12) | 0.257 |
| VO2max(L/min) | 3.34 (0.94) | 3.45 (0.93) | 0.135 |
| VEmax (L/min) | 125 (38) | 131 (37) | 0.150 |
| RER_max | 1.18 (0.07) | 1.19 (0.07) | 0.201 |
| Workload at Termination (W/kg) | 3.79 (0.89) | 3.82 (0.95) | 0.723 |
| End of Test | 17.8 (3.6) | 17.8 (3.7) | 0.969 |
| Fatigue Index | 0.02 (0.13) (N=104) | 0.03 (0.12) (N=377) | 0.679 |
| Average Power (W) | 540 (147) | 557 (137) | 0.366 |
| Relative Average Power (per Body Mass) (W·kg⁻¹) | 7.09 (1.25) | 7.23 (1.23) | 0.490 |
| Maximum Power (W) | 689 (196) | 704 (183) | 0.501 |
| Relative Maximum Power (per Body Mass) (W·kg⁻¹) | 8.95 (1.43) | 9.10 (1.50) | 0.305 |
| Time to Reach Pmax (s) | 6.50 (2.47) | 6.63 (2.69) | 0.413 |
| Maximum Rotational Speed (rev·s⁻¹) | 2.04 (0.31) | 2.07 (0.35) | 0.348 |
| Strength-Speed Index | 85 (39) | 87 (36) | 0.503 |

[1]n / N (%); [2]Pearson's Chi-squared test; Kruskal-Wallis rank sum test; WBC, white blood cell count ($10^9·L^{-1}$); RBC, red blood cell count ($10^{12}·L^{-1}$); HGB, haemoglobin ($g·dL^{-1}$); HCT, haematocrit (%); MCV, mean corpuscular volume (fL); MCH, mean corpuscular haemoglobin (pg); MCHC, mean corpuscular haemoglobin concentration ($g·dL^{-1}$); PLT, platelets ($10^9·L^{-1}$); TC, total cholesterol ($mg·dL^{-1}$); HDL, high-density lipoprotein cholesterol ($mg·dL^{-1}$); LDL, low-density lipoprotein cholesterol ($mg·dL^{-1}$); TG, triglycerides ($mg·dL^{-1}$); HR, heart rate ($beats·min^{-1}$); HRmax, maximal heart rate ($beats·min^{-1}$); $VO_2max$, maximal oxygen uptake reported as $mL·kg^{-1}·min^{-1}$ (mass-normalized) and $L·min^{-1}$ (absolute); VEmax, maximal minute ventilation ($L·min^{-1}$); RER_max, maximal respiratory exchange ratio; Workload at Termination, final incremental load ($W·kg^{-1}$); Stage/Time of Test Termination, stage number and elapsed time (min) at cessation of the incremental test; End of Test, rating of perceived exertion (Borg 6–20); Fatigue Index, Wingate-derived fatigue metric; Average Power

(W) and Relative Average Power ($W \cdot kg^{-1}$); Maximum Power (W) and Relative Maximum Power ($W \cdot kg^{-1}$); Time to Reach Pmax, time to peak power (s); Maximum Rotational Speed ($rev \cdot s^{-1}$).

Table 6_SupplInfo. IL15 rs1589241 (N=501)

| **Characteristic** | | **CC** N = 307[1] | **CT** N = 166[1] | **TT** N = 28[1] | **P**[2] |
|---|---|---|---|---|---|
| Glucose ($mg \cdot dL^{-1}$) | | 95 (9) | 95 (10) | 96 (10) | 0.769 |
| Creatinine ($mg \cdot dL^{-1}$) | | 0.89 (0.15) (N=305) | 0.91 (0.14) | 0.92 (0.14) | 0.424 |
| Iron ($\mu g \cdot dL^{-1}$) | | 117 (47) (N=305) | 116 (46) | 107 (46) | 0.630 |
| WBC ($10^9 \cdot L^{-1}$) | | 5.62 (1.56) | 5.66 (1.54) | 5.83 (1.84) | 0.868 |
| RBC ($10^{12} \cdot L^{-1}$) | | 4.91 (0.42) | 4.88 (0.42) | 4.95 (0.45) | 0.591 |
| HGB ($g \cdot dL^{-1}$) | | 14.74 (1.24) | 14.68 (1.20) | 14.77 (1.45) | 0.630 |
| HCT (%) | | 43.64 (3.20) | 43.55 (3.02) | 43.88 (3.68) | 0.690 |
| MCV (fL) | | 89.0 (4.0) | 89.5 (3.8) | 88.9 (3.4) | 0.466 |
| MCH (pg) | | 30.06 (1.50) | 30.13 (1.42) | 29.89 (1.43) | 0.830 |
| MCHC ($g \cdot dL^{-1}$) | | 33.76 (0.86) | 33.69 (0.97) | 33.63 (0.97) | 0.464 |
| PLT ($10^9 \cdot L^{-1}$) | | 236 (49) | 232 (46) (N=165) | 230 (60) | 0.548 |
| TC ($mg \cdot dL^{-1}$) | | 190 (37) (N=305) | 191 (35) | 184 (31) | 0.519 |
| HDL ($mg \cdot dL^{-1}$) | | 67 (18) (N=306) | 67 (18) | 69 (21) | 0.870 |
| LDL ($mg \cdot dL^{-1}$) | | 117 (35) (N=306) | 118 (33) | 110 (34) | 0.529 |
| TG ($mg \cdot dL^{-1}$) | | 98 (55) (N=305) | 105 (73) | 97 (51) | 0.909 |
| Cortisol ($\mu g \cdot dL^{-1}$) | | 16.1 (5.4) (N=306) | 15.7 (5.2) | 15.8 (6.1) | 0.513 |
| $VO_2max$ (mL/min/kg) | LF | 119 / 307 (39%) | 60 / 166 (36%) | 11 / 28 (39%) | 0.845 |
| | HF | 188 / 307 (61%) | 106 / 166 (64%) | 17 / 28 (61%) | |
| Body Mass (kg) | | 77 (15) | 77 (15) | 75 (14) | 0.771 |
| Waist Circumference (cm) | | 84 (12) | 85 (12) | 83 (10) | 0.660 |
| Hip Circumference (cm) | | 98 (7) | 99 (7) | 98 (7) | 0.699 |
| BMI ($kg \cdot m^{-2}$) | | 24.5 (3.8) | 24.7 (3.7) | 24.1 (3.3) | 0.722 |
| HR ($beats \cdot min^{-1}$) | | 70 (13) (N=306) | 69 (12) | 70 (13) | 0.540 |
| Sex | Females | 98 / 307 (32%) | 56 / 166 (34%) | 8 / 28 (29%) | 0.838 |
| | Males | 209 / 307 (68%) | 110 / 166 (66%) | 20 / 28 (71%) | |
| Age (years) | | 36 (8) | 36 (8) | 34 (7) | 0.271 |
| Stage of Test Termination | | 5.65 (1.87) | 5.54 (1.87) | 5.86 (1.80) | 0.735 |

| | | | | |
|---|---|---|---|---|
| Time of Test Termination (min) | 14.9 (3.7) | 14.6 (3.7) | 15.3 (3.6) | 0.700 |
| HRmax (beats·min⁻¹) | 180 (12) | 179 (13) | 181 (12) | 0.615 |
| VO2max(mL/min/kg) | 46 (12) | 45 (12) | 45 (13) | 0.831 |
| VO2max(L/min) | 3.43 (0.93) | 3.43 (0.91) | 3.34 (1.05) | 0.943 |
| VEmax (L/min) | 130 (38) | 130 (37) | 125 (26) | 0.794 |
| RER_max | 1.19 (0.07) | 1.19 (0.06) | 1.21 (0.08) | 0.630 |
| Workload at Termination (W/kg) | 3.83 (0.94) | 3.78 (0.94) | 3.93 (0.90) | 0.781 |
| End of Test | 17.9 (3.7) | 17.6 (3.7) | 18.3 (3.6) | 0.700 |
| Fatigue Index | 0.03 (0.13) (N=296) | 0.02 (0.10) (N=157) | -0.02 (0.10) | 0.220 |
| Average Power (W) | 555 (140) | 548 (141) | 557 (130) | 0.847 |
| Relative Average Power (per Body Mass) (W·kg⁻¹) | 7.24 (1.22) | 7.09 (1.30) | 7.40 (0.93) | 0.517 |
| Maximum Power (W) | 705 (186) | 694 (189) | 692 (164) | 0.883 |
| Relative Maximum Power (per Body Mass) (W·kg⁻¹) | 9.13 (1.43) | 8.93 (1.62) | 9.19 (1.19) | 0.542 |
| Time to Reach Pmax (s) | 6.51 (2.10) | 6.85 (3.52) | 6.21 (1.71) | 0.703 |
| Maximum Rotational Speed (rev·s⁻¹) | 2.07 (0.34) | 2.03 (0.36) | 2.08 (0.27) | 0.602 |
| Strength-Speed Index | 86 (36) | 87 (40) | 84 (27) | 0.988 |

[1]n / N (%); [2]Kruskal-Wallis rank sum test; Pearson's Chi-squared test; WBC, white blood cell count ($10^9$·L⁻¹); RBC, red blood cell count ($10^{12}$·L⁻¹); HGB, haemoglobin (g·dL⁻¹); HCT, haematocrit (%); MCV, mean corpuscular volume (fL); MCH, mean corpuscular haemoglobin (pg); MCHC, mean corpuscular haemoglobin concentration (g·dL⁻¹); PLT, platelets ($10^9$·L⁻¹); TC, total cholesterol (mg·dL⁻¹); HDL, high-density lipoprotein cholesterol (mg·dL⁻¹); LDL, low-density lipoprotein cholesterol (mg·dL⁻¹); TG, triglycerides (mg·dL⁻¹); HR, heart rate (beats·min⁻¹); HRmax, maximal heart rate (beats·min⁻¹); $VO_2$max, maximal oxygen uptake reported as mL·kg⁻¹·min⁻¹ (mass-normalized) and L·min⁻¹ (absolute); VEmax, maximal minute ventilation (L·min⁻¹); RER_max, maximal respiratory exchange ratio; Workload at Termination, final incremental load (W·kg⁻¹); Stage/Time of Test Termination, stage number and elapsed time (min) at cessation of the incremental test; End of Test, rating of perceived exertion (Borg 6–20); Fatigue Index, Wingate-derived fatigue metric; Average Power (W) and Relative Average Power (W·kg⁻¹); Maximum Power (W) and Relative Maximum Power (W·kg⁻¹); Time to Reach Pmax, time to peak power (s); Maximum Rotational Speed (rev·s⁻¹).

Table 7_SupplInfo. *TNFa* rs1800629 (N=501)

| **Characteristic** | **AA** N = 13[1] | **AG** N = 119[1] | **GG** N = 369[1] | **P**[2] |
|---|---|---|---|---|
| Glucose (mg·dL⁻¹) | 98 (18) | 96 (10) | 95 (9) | 0.876 |
| Creatinine (mg·dL⁻¹) | 0.81 (0.17) | 0.90 (0.13) (N=118) | 0.90 (0.15) (N=368) | 0.315 |

| | | | | | |
|---|---|---|---|---|---|
| Iron (µg·dL⁻¹) | | 111 (31) | 108 (47) (N=118) | 119 (47) (N=368) | 0.052 |
| WBC (10⁹·L⁻¹) | | 6.09 (1.93) | 5.50 (1.43) | 5.68 (1.60) | 0.550 |
| RBC (10¹²·L⁻¹) | | 4.96 (0.32) | 4.95 (0.40) | 4.88 (0.44) | 0.413 |
| HGB (g·dL⁻¹) | | 14.68 (1.03) | 14.76 (1.22) | 14.71 (1.25) | 0.906 |
| HCT (%) | | 43.77 (3.16) | 43.75 (3.02) | 43.58 (3.21) | 0.858 |
| MCV (fL) | | 88.3 (3.3) | 88.5 (4.1) | 89.4 (3.9) | 0.171 |
| MCH (pg) | | 29.62 (1.07) | 29.84 (1.63) | 30.16 (1.41) | 0.111 |
| MCHC (g·dL⁻¹) | | 33.55 (0.62) | 33.71 (0.97) | 33.74 (0.90) | 0.598 |
| PLT (10⁹·L⁻¹) | | 223 (50) | 233 (52) (N=118) | 235 (48) | 0.737 |
| TC (mg·dL⁻¹) | | 182 (26) | 195 (36) (N=118) | 189 (36) (N=368) | 0.080 |
| HDL (mg·dL⁻¹) | | 68 (18) | 66 (18) | 68 (18) (N=368) | 0.572 |
| LDL (mg·dL⁻¹) | | 107 (26) | 122 (35) | 115 (34) (N=368) | **0.037** |
| TG (mg·dL⁻¹) | | 103 (66) | 105 (77) (N=118) | 99 (56) (N=368) | 0.939 |
| Cortisol (µg·dL⁻¹) | | 15.8 (5.6) | 15.5 (4.5) | 16.1 (5.6) (N=368) | 0.845 |
| $VO_2$max (mL/min/kg) | LF | 4 / 13 (31%) | 60 / 119 (50%) | 126 / 369 (34%) | **0.005** |
| | HF | 9 / 13 (69%) | 59 / 119 (50%) | 243 / 369 (66%) | |
| Body Mass (kg) | | 77 (21) | 79 (15) | 76 (15) | 0.092 |
| Waist Circumference (cm) | | 85 (20) | 86 (12) | 83 (11) | **0.022** |
| Hip Circumference (cm) | | 100 (14) | 99 (7) | 98 (7) | 0.120 |
| BMI (kg·m⁻²) | | 25.1 (7.3) | 25.0 (3.5) | 24.4 (3.7) | 0.142 |
| HR (beats·min⁻¹) | | 68 (11) | 70 (12) | 70 (13) (N=368) | 0.836 |
| Sex | Females | 6 / 13 (46%) | 33 / 119 (28%) | 123 / 369 (33%) | 0.293 |
| | Males | 7 / 13 (54%) | 86 / 119 (72%) | 246 / 369 (67%) | |
| Age (years) | | 37 (9) | 35 (8) | 36 (8) | 0.273 |
| Stage of Test Termination | | 4.92 (2.10) | 5.41 (1.93) | 5.72 (1.83) | 0.136 |
| Time of Test Termination (min) | | 13.4 (4.2) | 14.5 (3.8) | 15.0 (3.6) | 0.192 |
| HRmax (beats·min⁻¹) | | 176 (15) | 181 (13) | 179 (12) | 0.218 |
| VO2max(mL/min/kg) | | 43 (13) | 44 (13) | 46 (12) | 0.367 |
| VO2max(L/min) | | 3.24 (1.07) | 3.43 (0.93) | 3.43 (0.92) | 0.764 |
| VEmax (L/min) | | 129 (43) | 129 (37) | 130 (37) | 0.879 |
| RER_max | | 1.21 (0.10) | 1.19 (0.07) | 1.19 (0.07) | 0.826 |

| | | | | |
|---|---|---|---|---|
| Workload at Termination (W/kg) | 3.46 (1.05) | 3.71 (0.96) | 3.86 (0.92) | 0.142 |
| End of Test | 16.4 (4.2) | 17.5 (3.8) | 18.0 (3.6) | 0.192 |
| Fatigue Index | 0.00 (0.07) (N=12) | 0.04 (0.11) (N=117) | 0.02 (0.12) (N=352) | 0.058 |
| Average Power (W) | 483 (178) | 566 (130) | 551 (141) | 0.271 |
| Relative Average Power (per Body Mass) (W·kg⁻¹) | 6.52 (2.00) | 7.15 (1.15) | 7.24 (1.23) | 0.314 |
| Maximum Power (W) | 678 (229) | 720 (182) | 695 (185) | 0.495 |
| Relative Maximum Power (per Body Mass) (W·kg⁻¹) | 8.69 (1.53) | 9.04 (1.48) | 9.09 (1.49) | 0.591 |
| Time to Reach Pmax (s) | 7.72 (5.08) | 6.49 (3.03) | 6.60 (2.38) | 0.052 |
| Maximum Rotational Speed (rev·s⁻¹) | 1.97 (0.35) | 2.06 (0.37) | 2.06 (0.33) | 0.607 |
| Strength-Speed Index | 85 (39) | 93 (41) | 84 (35) | 0.233 |

[1]n / N (%); [2]Kruskal-Wallis rank sum test; Pearson's Chi-squared test; WBC, white blood cell count ($10^9·L^{-1}$); RBC, red blood cell count ($10^{12}·L^{-1}$); HGB, haemoglobin ($g·dL^{-1}$); HCT, haematocrit (%); MCV, mean corpuscular volume (fL); MCH, mean corpuscular haemoglobin (pg); MCHC, mean corpuscular haemoglobin concentration ($g·dL^{-1}$); PLT, platelets ($10^9·L^{-1}$); TC, total cholesterol ($mg·dL^{-1}$); HDL, high-density lipoprotein cholesterol ($mg·dL^{-1}$); LDL, low-density lipoprotein cholesterol ($mg·dL^{-1}$); TG, triglycerides ($mg·dL^{-1}$); HR, heart rate ($beats·min^{-1}$); HRmax, maximal heart rate ($beats·min^{-1}$); $VO_2max$, maximal oxygen uptake reported as $mL·kg^{-1}·min^{-1}$ (mass-normalized) and $L·min^{-1}$ (absolute); VEmax, maximal minute ventilation ($L·min^{-1}$); RER_max, maximal respiratory exchange ratio; Workload at Termination, final incremental load ($W·kg^{-1}$); Stage/Time of Test Termination, stage number and elapsed time (min) at cessation of the incremental test; End of Test, rating of perceived exertion (Borg 6–20); Fatigue Index, Wingate-derived fatigue metric; Average Power (W) and Relative Average Power ($W·kg^{-1}$); Maximum Power (W) and Relative Maximum Power ($W·kg^{-1}$); Time to Reach Pmax, time to peak power (s); Maximum Rotational Speed ($rev·s^{-1}$).

Table 8_SupplInfo. *IL6* haplotype global (omnibus) associations for exploratory phenotypes

| Phenotype | P | FDR P |
|---|---|---|
| Glucose (mg·dL⁻¹) | 0.137 | 0.984 |
| Creatinine (mg·dL⁻¹) | 0.033 | 0.976 |
| WBC (10⁹·L⁻¹) | 0.365 | 0.986 |
| RBC (10¹²·L⁻¹) | 0.129 | 0.984 |
| HGB (g·dL⁻¹) | 0.082 | 0.984 |
| MCV (fL) | 0.986 | 0.986 |
| MCH (pg) | 0.983 | 0.986 |
| MCHC (g·dL⁻¹) | 0.385 | 0.986 |
| PLT (10⁹·L⁻¹) | 0.045 | 0.976 |
| TC_bad (mg·dL⁻¹) | 0.715 | 0.986 |

| | | |
|---|---|---|
| TC (mg·dL⁻¹) | 0.722 | 0.986 |
| HDL (mg·dL⁻¹) | 0.332 | 0.986 |
| LDL (mg·dL⁻¹) | 0.786 | 0.986 |
| TG (mg·dL⁻¹) | 0.768 | 0.986 |
| Cortisol (µg·dL⁻¹) | 0.902 | 0.986 |
| Body Mass (kg) | 0.642 | 0.986 |
| Waist Circumference (cm) | 0.536 | 0.986 |
| Hip Circumference (cm) | 0.780 | 0.986 |
| BMI (kg·m⁻²) | 0.798 | 0.986 |
| HR (beats·min⁻¹) | 0.642 | 0.986 |
| Stage of Test Termination | 0.966 | 0.986 |
| Time of Test Termination (min) | 0.907 | 0.986 |
| HRmax (beats·min⁻¹) | 0.342 | 0.986 |
| $VO_2$max (mL/min/kg) | 0.815 | 0.986 |
| $VO_2$max (L/min) | 0.657 | 0.986 |
| VEmax (L/min) | 0.108 | 0.984 |
| RER_max | 0.674 | 0.986 |
| Workload at Termination (W/kg) | 0.965 | 0.986 |
| End of Test | 0.907 | 0.986 |
| Mean | 0.701 | 0.986 |
| Minimum | 0.911 | 0.986 |
| Maximum | 0.778 | 0.986 |
| Fatigue Index | 0.767 | 0.986 |
| Average Power (W) | 0.279 | 0.986 |
| Relative Average Power (per Body Mass) (W·kg⁻¹) | 0.308 | 0.986 |
| Maximum Power (W) | 0.562 | 0.986 |
| Relative Maximum Power (per Body Mass) (W·kg⁻¹) | 0.519 | 0.986 |
| Time to Reach Pmax (s) | 0.822 | 0.986 |

| | | |
|---|---|---|
| Maximum Rotational Speed ($rev \cdot s^{-1}$) | 0.496 | 0.986 |
| Strength-Speed Index | 0.793 | 0.986 |

WBC, white blood cell count ($10^9 \cdot L^{-1}$); RBC, red blood cell count ($10^{12} \cdot L^{-1}$); HGB, haemoglobin ($g \cdot dL^{-1}$); HCT, haematocrit (%); MCV, mean corpuscular volume (fL); MCH, mean corpuscular haemoglobin (pg); MCHC, mean corpuscular haemoglobin concentration ($g \cdot dL^{-1}$); PLT, platelets ($10^9 \cdot L^{-1}$); TC, total cholesterol ($mg \cdot dL^{-1}$); HDL, high-density lipoprotein cholesterol ($mg \cdot dL^{-1}$); LDL, low-density lipoprotein cholesterol ($mg \cdot dL^{-1}$); TG, triglycerides ($mg \cdot dL^{-1}$); HR, heart rate ($beats \cdot min^{-1}$); HRmax, maximal heart rate ($beats \cdot min^{-1}$); $VO_2max$, maximal oxygen uptake reported as $mL \cdot kg^{-1} \cdot min^{-1}$ (mass-normalized) and $L \cdot min^{-1}$ (absolute); VEmax, maximal minute ventilation ($L \cdot min^{-1}$); RER_max, maximal respiratory exchange ratio; Workload at Termination, final incremental load ($W \cdot kg^{-1}$); Stage/Time of Test Termination, stage number and elapsed time (min) at cessation of the incremental test; End of Test, rating of perceived exertion (Borg 6–20); Fatigue Index, Wingate-derived fatigue metric; Average Power (W) and Relative Average Power ($W \cdot kg^{-1}$); Maximum Power (W) and Relative Maximum Power ($W \cdot kg^{-1}$); Time to Reach Pmax, time to peak power (s); Maximum Rotational Speed ($rev \cdot s^{-1}$).

Table 9_SupplInfo. Term-wise P values for *IL6×IL15* and *IL6×TNFα* interaction coefficients across phenotypes.

| Phenotype | *IL6×IL15* | *IL6×TNFα* |
|---|---|---|
| Iron | 0.784 | 0.775 |
| HCT | 0.482 | 0.113 |
| LDL | 0.504 | 0.743 |
| $VO_2max$ HF vs LF | 0.526 | 0.294 |

Likelihood ratios test comparing the models with and without an interaction term, adjusted for age and sex. HCT, haematocrit; LDL, low-density lipoprotein cholesterol; $VO_2max$, maximal oxygen uptake; HF, High Fitness; LF, Low Fitness. $P < 0.05$ would indicate a significant interaction; no significant interactions were observed.
